# *TREM2* Risk Variants with Alzheimer’s Disease Differ in Rate of Cognitive Decline

**DOI:** 10.1101/2024.10.17.24315643

**Authors:** Janna I.R. Dijkstra, Lisa Vermunt, Vikram Venkatraghavan, Georgii Ozgehov, Emma M. Coomans, Rik Ossenkoppele, Elsmarieke van de Giessen, Marc Hulsman, Christa M. de Geus, Wiesje M. van der Flier, Sietske A.M. Sikkes, Frederik Barkhof, Betty Tijms, Alida A. Gouw, Willem de Haan, Everard G.B. Vijverberg, Yolande A.L. Pijnenburg, Henne Holstege, Charlotte E. Teunissen, Sven J. van der Lee

## Abstract

Rare variants of the triggering receptor expressed on myeloid cell 2 (*TREM2*) gene are major risk factors for Alzheimer’s disease (AD), and drugs targeting the TREM2 protein are being developed. However, it is unknown whether carriers of a *TREM2* risk variant have a clinically distinct AD phenotype. Here we studied a full range of clinical measures in a large cohort of *TREM2* variant carriers (*n*=123, 7.8%, i.e., R62H *n*=66, R47H *n*=26, T96K *n*=16, other *TREM2* variants *n*=17) compared to confirmed non-carriers (*n*=1,459) with biomarker confirmed symptomatic AD from Amsterdam Dementia Cohort.

*TREM2* variant carriers (mean age at diagnosis 64.4 years (SD ±7.1), 54% female) did not show distinct clinical measures of AD at presentation compared to AD patients not carrying a *TREM2* variant (mean age at diagnosis 64.4 ±7.0, 52% female). Specifically, we observed no differences in MMSE, most neuropsychological domains (except visuospatial functioning), MRI scores, CSF biomarkers, and EEG. Also, in an exploratory analysis of neuroimaging measures, including structural MRI (41 ROIs) and Tau-PET scans of four carriers (R62H, R47H, G58A, D87N), *TREM2* variant carriers showed similar atrophy patterns and similar abnormal tracer binding compared to non-carriers. Despite not being different at baseline, carriers did show faster cognitive decline in follow-up. Carriers declined 0.63 ±0.25 points on the MMSE more per year compared to non-carriers, but there was no difference in the hazard rate of death after diagnosis.

Finally, we explored whether specific *TREM2* variants are associated with distinct clinical measures compared to the reference group, i.e. non-carriers, within the same cohort. Notably, both R47H and T96K carriers exhibited faster cognitive decline, and R47H carriers even showed an increased rate of death after diagnosis. In contrast to the shared cognitive decline, these variants showed different results for other measures at baseline.

This study presents a detailed overview of the clinical measures in AD patients carrying a *TREM2* risk variant, and it shows that carriers of *TREM2* risk variants cannot be distinguished based on clinical presentation at baseline. However, carriers exhibit a faster global cognitive decline compared to non-carriers. Variant-specific analyses suggest that especially R47H and T96K carriers drive the association of *TREM2* variants with faster cognitive decline.

## MAIN

Rare *TREM2* variants are major risk factors for Alzheimer’s disease (AD) (1–6). The triggering receptor expressed on myeloid cell 2 (*TREM2*) gene is situated on chromosome 6, it encodes a transmembrane protein of 230 amino acids, and it is expressed exclusively in microglia within the brain (7). The TREM2 protein appears to be a key player in microglial function and AD development (8–10), and is a target of disease-modifying therapies that are currently in phase II clinical trials (11–16).

To date, it is unknown whether carriers of a *TREM2* risk variant have a specific clinical presentation of AD. In a retrospective study of autopsied cases, *TREM2* variant carriers more often had non-amnestic syndromes compared to non-carriers, faster cognitive decline (17), more tau accumulation, but no altered regional beta-amyloid (Aβ) burden (17,18). Another study did not find a distinct neuropsychological profile when comparing *TREM2* R47H carriers with AD non-carriers (19). All these studies were small with a maximum number of 31 *TREM2* variant carriers. Therefore, the variability of results between studies may be explained by small samples (20,21) and heterogeneity of effects introduced by studying populations of different ancestry (4,21), both making it more difficult to find associations.

Another explanation why associations with clinical measures are inconclusive could be the variant-specific effects. At a molecular level, *TREM2* risk variants impair TREM2 activity differently (7,22–24). Most *TREM2* risk variants are situated on exon 2 where the coding corresponds to the Ig-like V type domain (25), suggesting an alteration in the interaction between TREM2 and its ligands (21,25). R47H is located near the exon 2 junction, whereas T96K is located near a conserved part of the protein; thus, these variants could affect distinct functional regions on TREM2’s surface (23). Several studies indicated that TREM2 proteins resulting from R47H showed reduced ligand binding and signalling, while conversely proteins resulting from T96K showed enhanced ligand binding (22,24). In addition, the variants R62H and R47H associated with two different AD subtypes based on CSF proteomics (26), which further indicates variant-specific mechanisms. Hence, *TREM2* variant-specific mechanisms necessitate variant-specific studies. Studying this hypothesis requires large clinical sample sizes to be able to observe adequate numbers for variant-specific analyses. Previous research indicated that *TREM2* R47H carriers seem to show a typical clinical AD profile (27), elevated CSF-Tau (28), and lower grey matter volume in right orbitofrontal regions compared to non-carriers (19). However, another study did not find a significant effect on cross-sectional brain volumes (29). *TREM2* R62H and T96K carriers have not yet been studied well.

Here we hypothesize that *TREM2* risk variants may be associated with distinct clinical measures. Hence, we studied the association of *TREM2* carriership with a full range of clinical measures at baseline (neuropsychological profile, visual MRI rating, CSF AD biomarkers, and visual EEG rating) and in follow-up (cognitive decline and survival status) in a large clinical cohort of biomarker confirmed AD patients, followed by an exploratory analysis of neuroimaging measures (structural MRI, and Tau-PET) and an analysis of the specific *TREM2* variants (R47H, R62H, T96K and others).

## METHODS

### Amsterdam Dementia Cohort

We included 1,582 patients with Mild Cognitive Impairment (MCI) or dementia due to AD, based on confirmed AD biomarkers (in CSF 95% and amyloid PET 5%), and with available genetic data who visited the Alzheimer Centre Amsterdam memory clinic (Fig. 1) (31). We identified a *TREM2* risk variant in 123 AD patients, representing 7.8% of the total cohort, while 1,459 AD patients were confirmed to not carry a *TREM2* risk variant. All patients underwent a standardized diagnostic trajectory (31). Information was collected on demographics, medical history, family history, neuropsychological investigation, MRI, cerebrospinal fluid (CSF), and blood. Diagnoses were determined by consensus in a multidisciplinary meeting, ensuring that diagnostic criteria were met. Patients were followed over time for reassessments and/or research purposes. Patients with a revised diagnosis at follow-up (*n*=22) were excluded. Mortality data was collected from the Central Public Administration. Patients consented to be part of the Amsterdam Dementia Cohort (ADC) to use their medical information for research and to allow their DNA to be stored in a dedicated biobank.

**Fig. 1:**
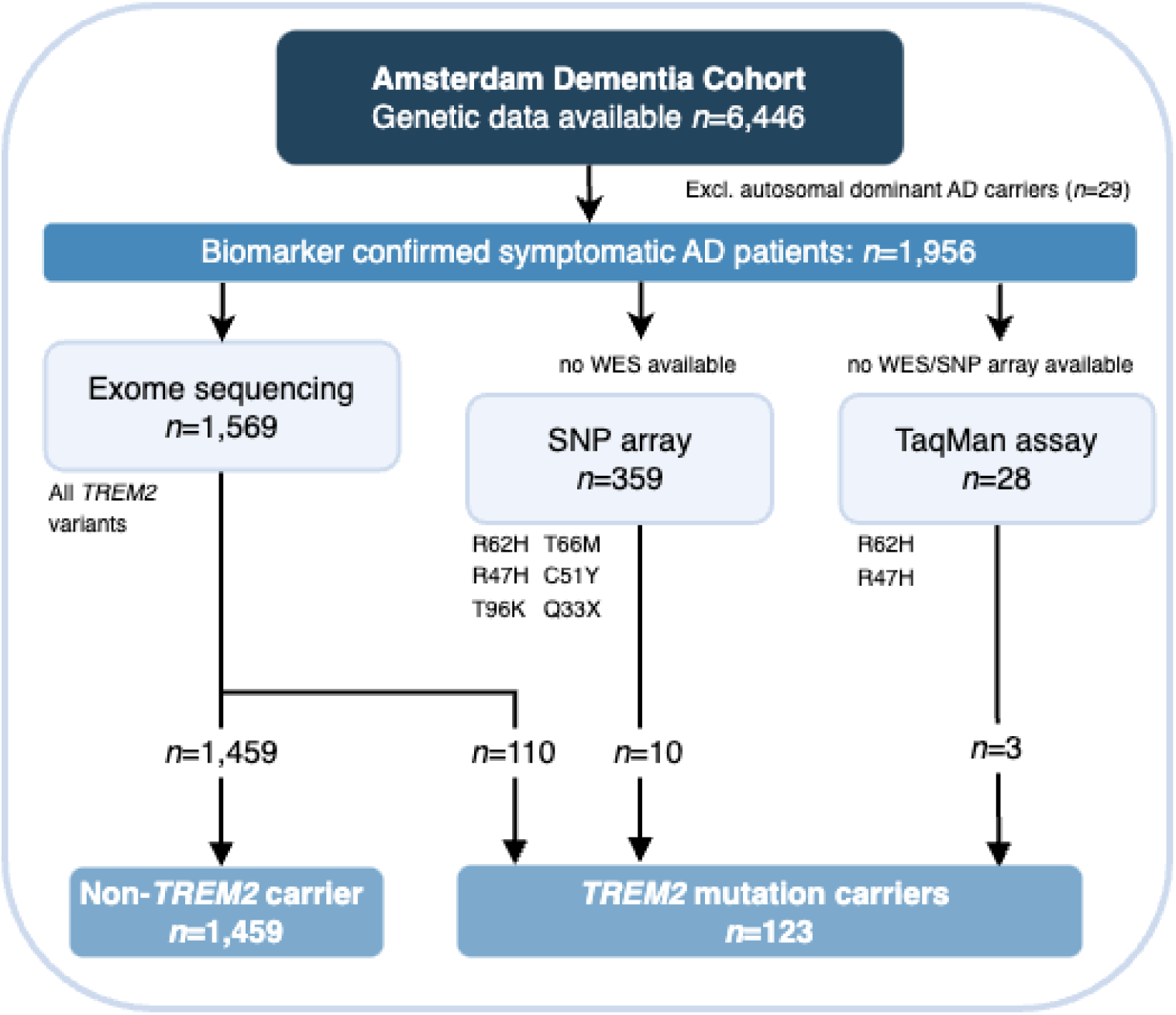
Flowchart of population from the Amsterdam Dementia Cohort. Patients were included from the Amsterdam Dementia Cohort. *TREM2* carriership was determined by whole exome sequencing (WES), SNP array, and targeted TaqMan assay (including *TREM2* variants R47H and R62H); and *TREM2* non-carriership was determined by WES. Patients were included with an amyloid confirmed diagnosis of mild cognitive impairment (MCI) or dementia based on Alzheimer’s disease. Abbreviations: A105V = p.Ala105Val; AD = Alzheimer’s disease; ADAD = Autosomal dominant Alzheimer’s disease; C51Y = p.Cys51Tyr; CSF = Cerebrospinal Fluid; D87N = p.Asp87Asn; G58A = p.Gly58Ala; MRI = Magnetic Resonance Imaging; SNP = Single-Nucleotide Polymorphism; Q33X = p.Gln33Ter; R31F = p.Ser31Phe; R47G = p.Arg47Gly; R47H = p.Arg47His; R62H = p.Arg62His; T96K = p.Thr96Lys; *TREM2* = Triggering Receptor Expressed on Myeloid Cells 2; WES = Whole Exome Sequencing.

### Genotyping, Imputation of 6 Selected SNPs, and Genetic Selection

*Whole exome sequencing, Single-Nucleotide Polymorphism (SNP) Arrays and targeted TaqMan assay* After DNA collection, most samples were genotyped with whole exome sequencing (WES) data using Illumina sequencers (*n*=1,569 of total 1,956). Some samples did not have WES but were genotyped on Illumina Global Screening Array (*n*=359). We included six missense variants that were imputed with high quality (R^2^ >0.37) (R47H, R62H, T96K, T66M, C51Y, and Q33X) (32). Standard quality control was performed. Samples were imputed with the Trans-Omics for Precision Medicine (TOPMeD) reference panel (33,34). Processing, quality control and variant calling has been described previously (3,35). For patients without WES or SNP array data available, made-to-order TaqMan assays were targeted on variants R47H and R62H (*n*=28). Patients with autosomal dominant AD, i.e., carriers of genetic mutations in *PSEN1*, *PSEN2* and *APP*, were excluded (*n*=29).

#### Genetic selection

A schematic overview of the population is presented in Fig 1. Whole exome sequencing (WES) (*n*=110), SNP array (*n*=10), and TaqMan assays (*n*=3) identified 123 *TREM2* carriers, and WES confirmed that 1,459 AD patients did not carry a *TREM2* variant (i.e., non-carriers). We included *TREM2* missense variants proven to be associated with AD. These are R47H (NC_000006.12: g.41161515G>C, OR 3.1, p<0.5×10^e-5^) (1,6), R62H (NC_000006.12: g.41161470G>A, OR 1.7, p<0.5×10^e-5^) (6), and T96K (NC_000006.12: g.41129105 C>A, OR 1.2 in African GWAS, p<0.5×10^e-5^) (4). One single variant was selected based on biological evidence, being D87N (36). Other rare damaging variants, including splice variants, were selected based on the burden test association. This test included a Rare Exome Variant Ensemble Learner (REVEL) score >0.25 (3,37), protein truncating variants, or frameshift deletion variants.

### Clinical measures

#### Neuropsychological assessment

Global cognitive functioning was assessed using the Mini-Mental State Examination (MMSE) (38). MMSE data was available for 1,564 (99%) patients. In addition, neuropsychological data was available for 1,519 (96%) patients. We measured five neuropsychological domains, i.e., episodic memory, executive functioning, attention and speed, language, and visuospatial functioning using a standardized neuropsychological assessment comprising eight cognitive tests (31). Classification was based on a total of 16 variables as previously reported by Dubbelman et al. (2022) (39). Supplementary Data gives an overview of the variables used per domain. In short, each domain was assessed when at least two cognitive tests were available (range of available data per domain: 69-91%). Z-scores for each variable were calculated per cognitive domain scaled on the baseline mean and standard deviation of the total cohort. The combined domains gave a summarized z-score of global cognition. Longitudinally, we studied cognitive decline using the MMSE. MMSE data had a median follow-up of 1.0 years (interquartile range (IQR) 0.0-2.4); 42% had one measure, 21% had two measures, 15% had three measures, and 22% had more than three measures.

#### CSF biomarkers

CSF data was available for 1,522 (96%) patients. CSF Aβ42, pTau-181 and total tau were assessed with the Innotest enzyme-linked immunosorbent assay (ELISA) and Aβ42 was drift corrected, or on Elecsys (40). Amyloid status for AD was confirmed if the Innotest tau/Aβ42 ratio exceeded 0.46 (41), or the Elecsys pTau-181/Aβ42 ratio exceeded 0.20 (42). If an (additional) amyloid-PET scan was available, amyloid status was confirmed by positive amyloid-PET scan (*n*=85). To correct for variance between these assays, Elecsys results were converted based on established equations in biomarker associations (43). In addition, we employed available CSF-NfL measurements described in a previous paper defining reference values for the SIMOA NF-light assay (44). To calculate age-adjusted z-scores in CSF, we approached the reference range percentile formula with a linear model with outcome log2(NfL) and age as the predictor in the reference. This resulted in the following formula for calculating age-adjusted NfL z-scores in CSF: [log2(NfL)⍰−⍰(6.661⍰+⍰(age⍰×⍰ 0.045))] / 0.736.

#### MRI clinical ratings

Brain-MRI data was available for 1,210 (76%) patients. Three visual rating scales as used in clinical assessment were employed: Medial Temporal lobe Atrophy (MTA) (45), Posterior Cortical Atrophy (PCA) (46), and Fazekas score for white matter hyperintensities (47).

#### Electroencephalogram (EEG)

EEG data was available for 1,304 (82%) patients. Details of the acquisition, processing, and visual assessment of the EEG recordings have been described previously (48). The assessment was conducted utilizing a standardized severity scale (1 to 4), representing the spectrum from no abnormalities to severe abnormalities (49). We studied ‘normal’ versus ‘abnormal’ EEG scans. ‘Normal’ was defined as 1-2 on the severity scale, with or without focal abnormalities. ‘Abnormal’ were all other possibilities, including epileptiform activity and diffuse abnormalities.

### Neuroimaging measures

#### MRI structural brain imaging

Structural MRI data was available for 1,069 (67%) patients. Quantitative image analysis was done for several regions of interest based on Desikan Kiliany Atlas by FreeSurfer v7.1 (50). Details of the MRI data processing (51) and quality check process (52) have been described previously. MRI data were from 12 scanners. The scanner-related effects were removed using a procedure called ComBat (53). We studied 34 cortical thickness measures (mm) and seven subcortical volumes (mm^3^). We averaged measures of the left and right hemisphere per region.

#### [^18^F]flortaucipir PET

Tau-PET data was available for 67 (4%) patients, including four *TREM2* variant carriers (R47H, R62H, G58A, and D87N). Details on the acquisition and processing of the [^18^F]flortaucipir PET images have been described previously (54–57). For semi-quantification, we calculated standardized uptake value ratio (SUVr) using the gray matter cerebellum as reference region in six different composite regions of interest from the Hammers and Svarer templates: 1) a medial temporal region (including the entorhinal cortex, parahippocampal gyrus, amygdala, and fusiform gyrus), 2) a lateral temporal region (including the superior, middle and inferior temporal gyrus, and the posterior temporal lobe), 3) a medial parietal region (including the posterior cingulate), 4) a lateral parietal region (including the superior parietal gyrus and the inferolateral remainder of the parietal lobe), 5) an occipital region (including the cuneus, lingual gyrus and lateral remainder of the occipital lobe), and 6) a frontal region (including the superior, middle and inferior frontal gyrus, gyrus rectus, and orbitofrontal gyrus). In line with previous work (58), we contrasted the SUVRs for the AD cases with a *TREM2* variant against the observed distribution of the AD cases without a TREM2 variant. Hippocampi could not be adequately assessed with this tracer due to off-target binding.

### Statistical analyses

Analyses were performed in R version 4.3.0 (59) and Python version 3.9 (60). Education level was converted from the Dutch Verhage scale (61) to the Standard Classification of Education, i.e., low, medium, and high (62). For individuals with missing education level (*n*=7), the median level was imputed, i.e., high education. Baseline characteristics of *TREM2* variant carriers and non-carriers were compared with Chi-squared tests for categorical variables, and with t-tests for continuous variables.

In the main analysis, all *TREM2* variants were combined to study the effect of *TREM2* mutation status with clinical measures relative to the reference group, i.e., non-carriers. Linear regression models were used to associate *TREM2* mutation status with MMSE at baseline, the five neuropsychological domains and combined global cognition, MRI features, and CSF biomarkers. All measurements were scaled for comparability. Logistic regression models were used to associate *TREM2* mutation status with an EEG abnormality score. Regression analyses were conducted separately and were adjusted for age and sex as dependent variables, and MMSE at baseline, neuropsychological profiles and EEG scores were also adjusted for education level (i.e., low, middle, high) (62) and disease stage (i.e., MCI or dementia). With Cox proportional hazards models, we associated *TREM2* mutation status with time in years between diagnosis and death adjusted for age at diagnosis, sex, education level, disease stage, and MMSE at baseline. Linear mixed models were used to study associations of *TREM2* variants and change in MMSE scores over time, and were adjusted for age at diagnosis, sex, education level, and disease stage. The models included a random intercept and interaction effect over time. A p-value corrected for a false discovery rate (FDR) <0.10 was considered statistically significant.

In the exploratory analysis of neuroimaging outcomes, we tested with linear regression models the effect of *TREM2* status with 41 regions of interest (ROI) from structural MRI, and models were adjusted for age, sex, and estimated intracranial volume.

In the subsequent exploratory analysis, *TREM2* variants were grouped to associate variant-specific *TREM2* effects with each clinical measure relative to the reference group, comparing (i) R62H carriers vs. non-carriers, (ii) R47H carriers vs. non-carriers, (iii) T96K carriers vs. non-carriers, and (iv) other *TREM2* carriers vs. non-carriers. In the cox regression models and linear mixed models, we tested a separate *TREM2* effect relative to the reference group comparing categorical (R62H, R47H, T96K, other) carriers vs. non-carriers. Carriers of two different mutations were categorized for the variant conferring the highest risk. A p-value <0.05 was considered statistically significant.

In the sensitivity analysis, we excluded patients of non-European descent (determined through 1000 Genomes clustering) (63) and patients who had a familial relationship (identity-by-descent ≥ 0.2). All the models described above were further adjusted for three principal components.

## RESULTS

### Population and *TREM2* characteristics

Ten different *TREM2* risk variants were identified (Table 1). All genetic variants were located on exon 2 (of 5 exons). The most prevalent mutations were R62H (*n*=66, 54%), R47H (*n*=26, 21%; among which one also carried R62H) and T96K (*n*=16, 13%, among which two were homozygous). We further observed rare mutations in seventeen patients (14% of *TREM2* carriers) that carried one of the following heterozygous missense, splice, or protein truncating mutations: p.Ser31Phe (R31F), p.Gln33Ter (Q33X; among which one also carrier R62H), p.Arg47Gly (R47G), p.Cys51Tyr (C51Y), p.Gly58Ala (G58A), p.Asp87Asn (D87N), and p.Ala105Val (A105V).

**Table 1:**
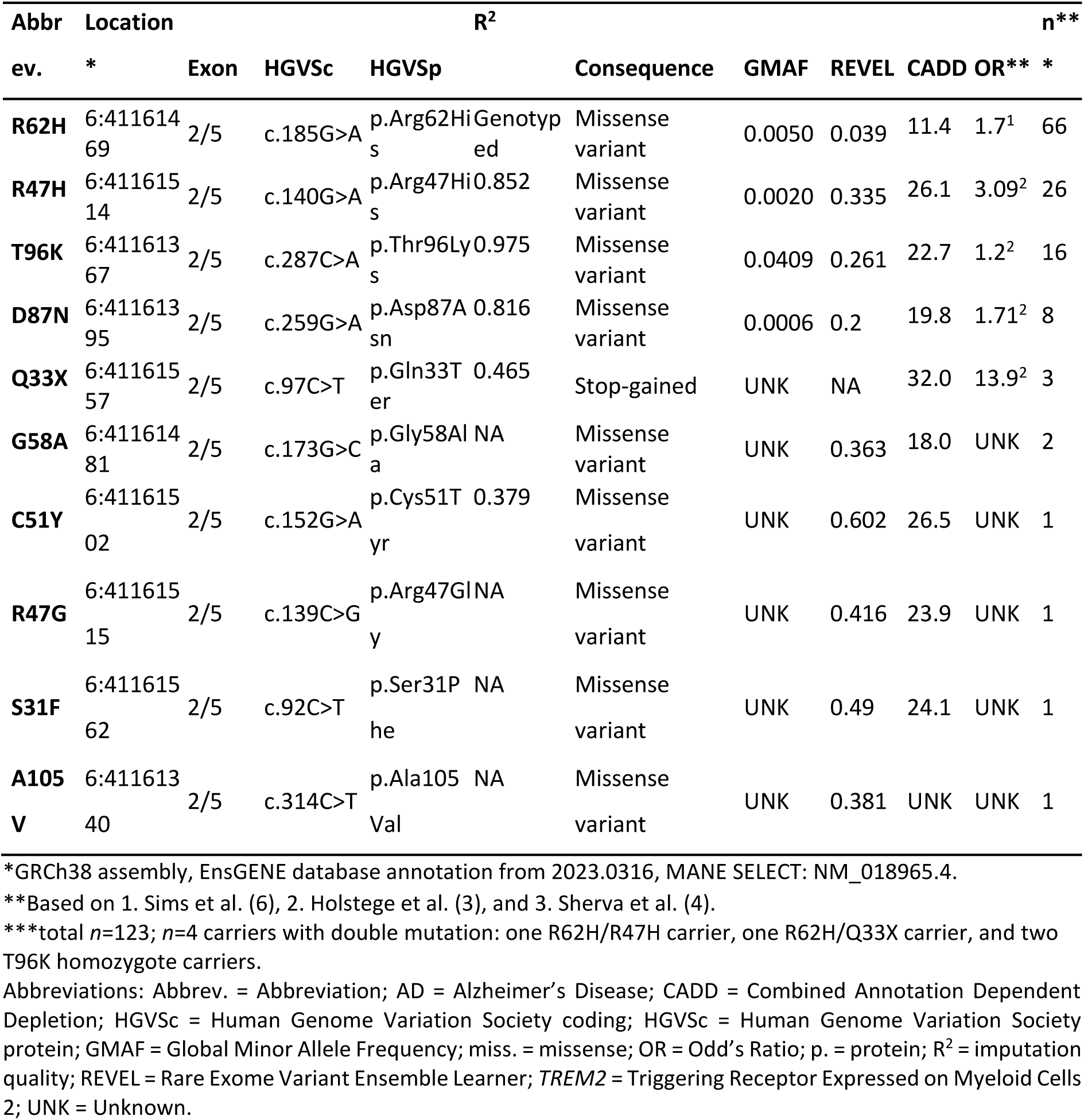
Genetic descriptives of *TREM2* variants identified in the cohort at baseline.

### Cohort demographics and data availability per clinical measure

*TREM2* variant carriers had a mean age at diagnosis of 64.4 years (standard deviation (SD) ±7.1), 67 were female (54%), and 71 died (58%) with a mean age at death of 70.4 ±7.9 years (Table 2). Non-carriers had a mean age at diagnosis of 64.4 ±7.0 years, 757 were female (52%), and 878 died (60%) with a mean age at death of 71.1 ±7.7 years. In addition, 57% of *TREM2* variant carriers had a positive family history (i.e., having an affected first-degree relative) compared to 45% of non-carriers (Chi-squared (X^2^) *P*=2.1×10^−2^), and 71% of *TREM2* carriers carried an *APOE-*ε*4* allele compared to 69% of non-carriers (χ^2^ *P*=0.71). Furthermore, 82% of *TREM2* variant carriers were diagnosed with dementia and 18% with MCI, compared to 87% of non-carriers with dementia and 13% with MCI (χ^2^ *P*=0.).

**Table 2:**
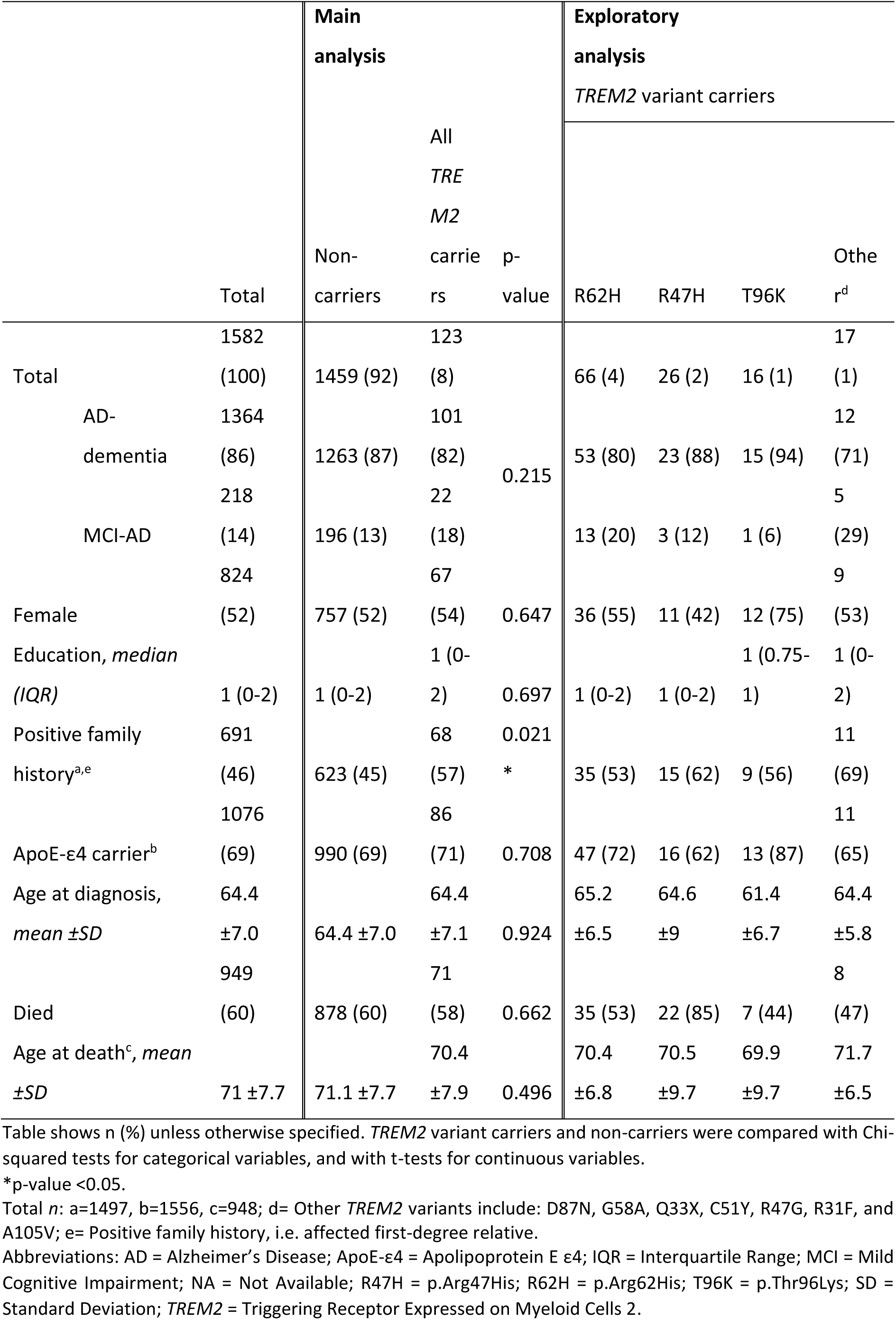
Demographics of cohort at baseline stratified by *TREM2* variant carriership.

### Main analysis

#### Effects of all TREM2 variants combined on clinical outcomes

Fig. 2 shows a heatmap of all outcomes from the main analysis and Fig. 3 summarizes all findings. *TREM2* variant carriers did not associate with MMSE at baseline or most neuropsychological domains compared to AD patients not carrying a *TREM2* variant, however they did show less impaired scores in visuospatial functioning (standardized *β* (std*β*) 0.21, ± standard error (se) 0.08, *P_fdr_* =9.4×10^−2^). *TREM2* carriership did not associate with MRI clinical ratings or CSF AD biomarker levels. In the longitudinal analysis, *TREM2* carriers showed a faster cognitive decline compared to non-carriers; non-carriers declined on average 1.80 points on MMSE per year of follow-up, whereas *TREM2* carriers declined on average 2.43 points (*β*-difference −0.63 ±0.25, *P_fdr_* =9.4×10^−2^) (Table 3, Fig. 5A). *TREM2* carriers were not at increased risk of mortality (Hazard Ratio (HR) 1.12, 95% Confidence Interval (CI) 0.9-1.4, *P_fdr_*=0.667) (Supplementary Table 3, Supplementary Fig. 1).

**Fig. 2:**
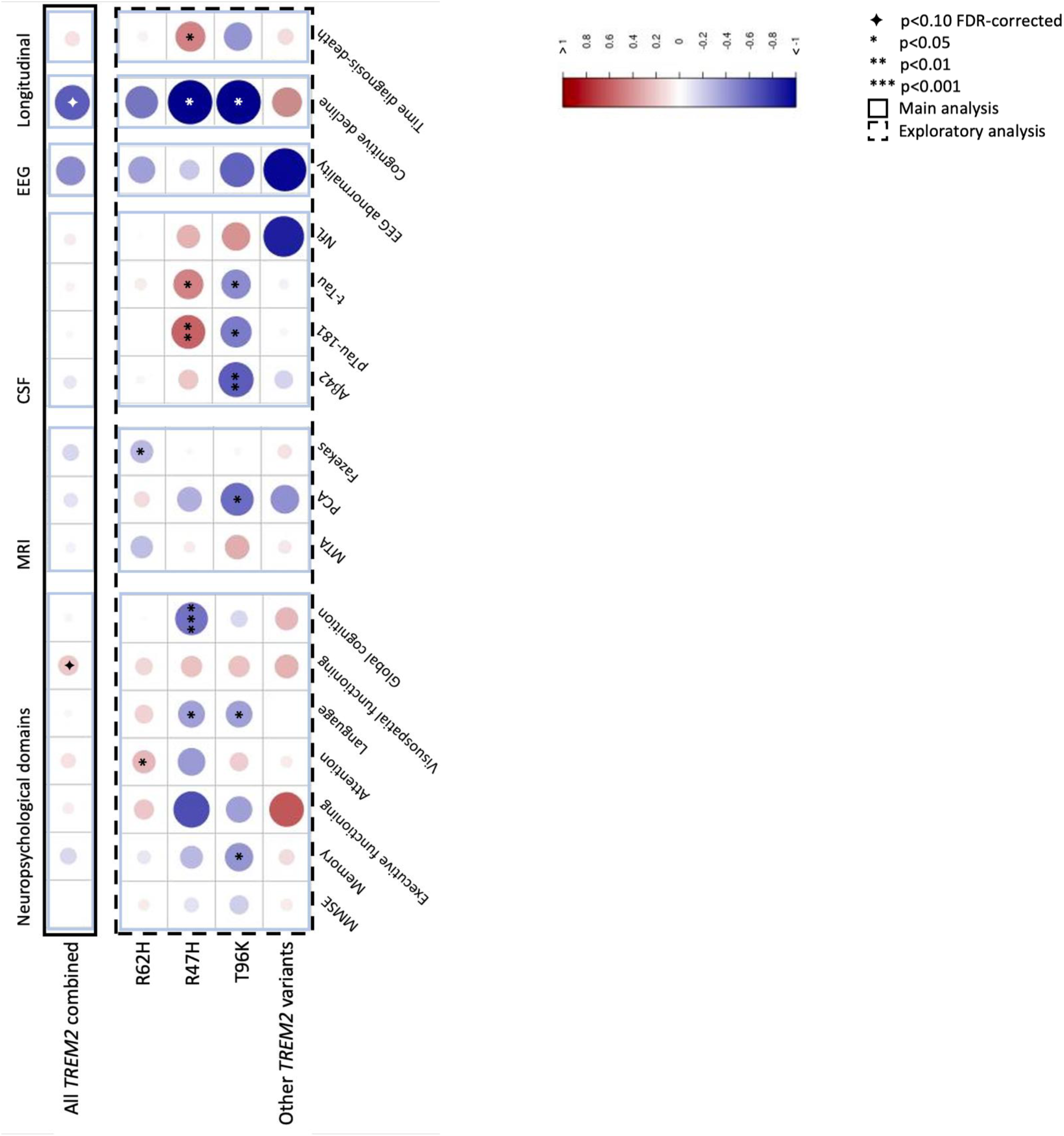
*TREM2* effect on AD clinical measures comparing *TREM2* risk variant carriers vs. non-carriers. Shown here are *standardized* betas of linear regression models adjusted for age and sex; MMSE at baseline and neuropsychological domains are also adjusted for education level and disease stage. Also shown here are betas of logistic regression models for EEG, cox regression models (Cox), and linear mixed models (LMM) adjusted for age, sex, education level, and disease stage; cox regression models are also adjusted for MMSE at baseline. Other *TREM2* variants include: D87N, G58A, Q33X, C51Y, R47G, R31F, and A105V. Abbreviations: Aβ42 = Beta-Amyloid 42; AD = Alzheimer’s Disease; CSF = Cerebrospinal Fluid; Cox = Cox regression models; EEG = Electroencephalogram; FDR = False Discovery Rate; LMM = Linear Mixed Models; MMSE = Mini-Mental State Examination; pTau-181 = phosphorylated Tau-181; R47H = p.Arg47His; R62H = p.Arg62His; T96K = p.Thr96Lys; *TREM2* = Triggering Receptor Expressed on Myeloid Cells 2.

**Fig. 3:**
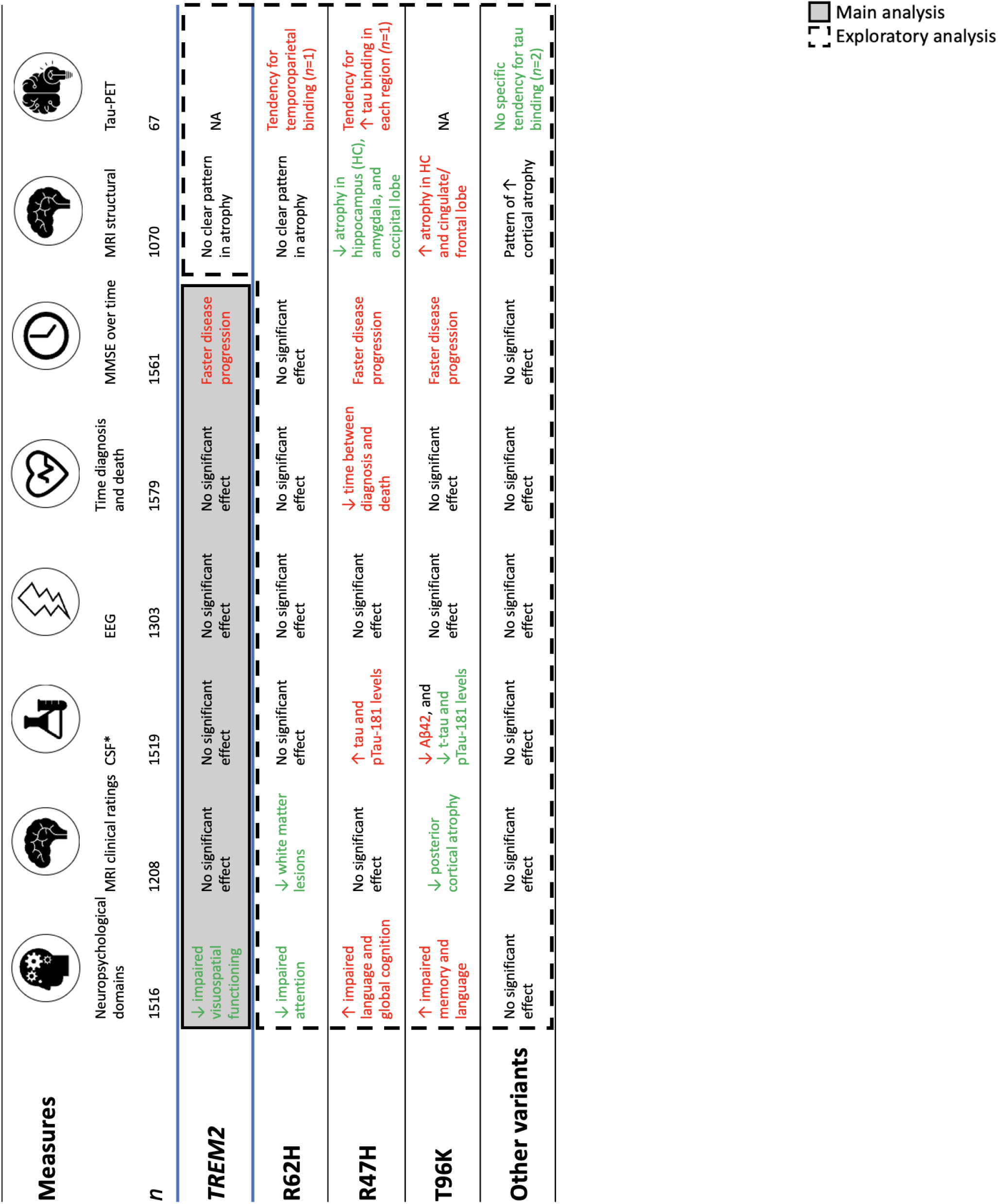
Main findings in this study. * CSF-NfL total *n*=165, *TREM2* mutation carriers=10: R62H=4, R47H=3, T96K=2, other variants=1. Other *TREM2* variants include: D87N, G58A, Q33X, C51Y, R47G, R31F, and A105V. Abbreviations: Aβ42 = Beta-Amyloid 42; CSF = Cerebrospinal fluid; MMSE = Mini-Mental State Examination; MRI = Magnetic Resonance Imaging; NA = Not Available; pTau-181: Phosphorylated Tau-181; *TREM2* = Triggering Receptor Expressed on Myeloid Cells 2.

**Table 3:**
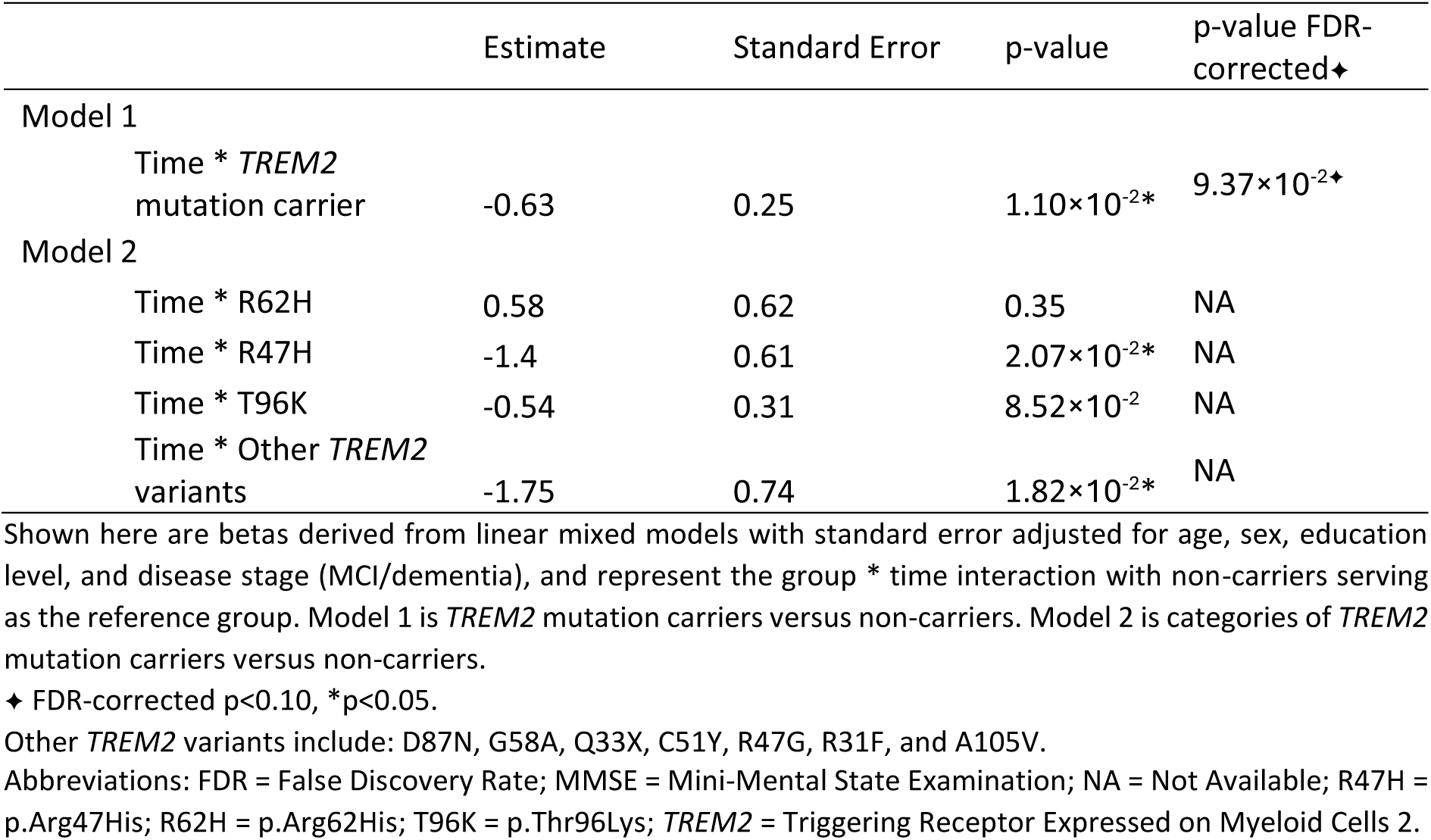
Effect of *TREM2* variants on cognitive decline (in MMSE) stratified by *TREM2* mutation carriers compared to non-carriers.

### Exploratory analysis of neuroimaging measures

#### Effects of TREM2 variants combined on structural MRI and tau-PET imaging

On structural MRI, *TREM2* variant carriers had smaller amygdala (std*β* 0.19 ±0.10, *P*=4.7×10^−2^) compared to AD patients not carrying a *TREM2* variant (Fig. 5). There was no difference in 34 cortical thickness regions or the six other subcortical volumes. Fig. 6A shows the tau-PET scans of each of the *TREM2* variant carriers as well as the average tau-PET scan of the non-carrier group. On visual inspection, each of the *TREM2* carriers showed clear increased tracer binding in temporoparietal regions, similarly to the average non-carrier AD group.

**Fig. 4:**
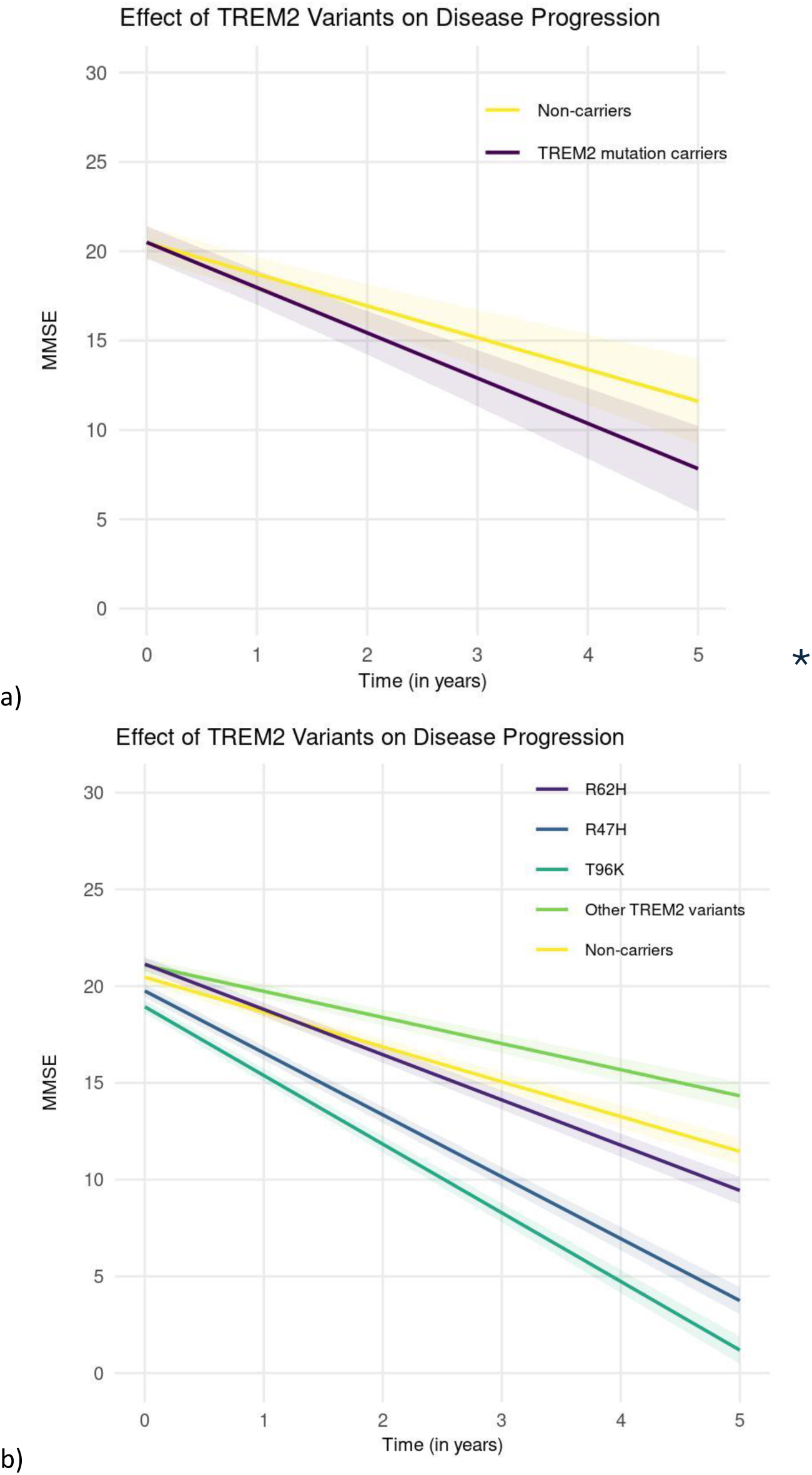
Effect of *TREM2* variants on cognitive decline in symptomatic AD patients compared to non-carriers. Shown here are linear mixed models with 95% confidence interval, adjusted for age, sex, education level, disease stage (MCI/dementia). (a) *TREM2* mutation carriers vs. non-carriers. (b) *TREM2* mutation carriers stratified by mutations vs. non-carriers. Other *TREM2* variants include: D87N, G58A, Q33X, C51Y, R47G, R31F, and A105V. Abbreviations: MMSE = Mini-Mental State Examination; R47H = p.Arg47His; R62H = p.Arg62His; T96K = p.Thr96Lys; *TREM2* = Triggering Receptor Expressed on Myeloid Cells 2.

**Fig. 5:**
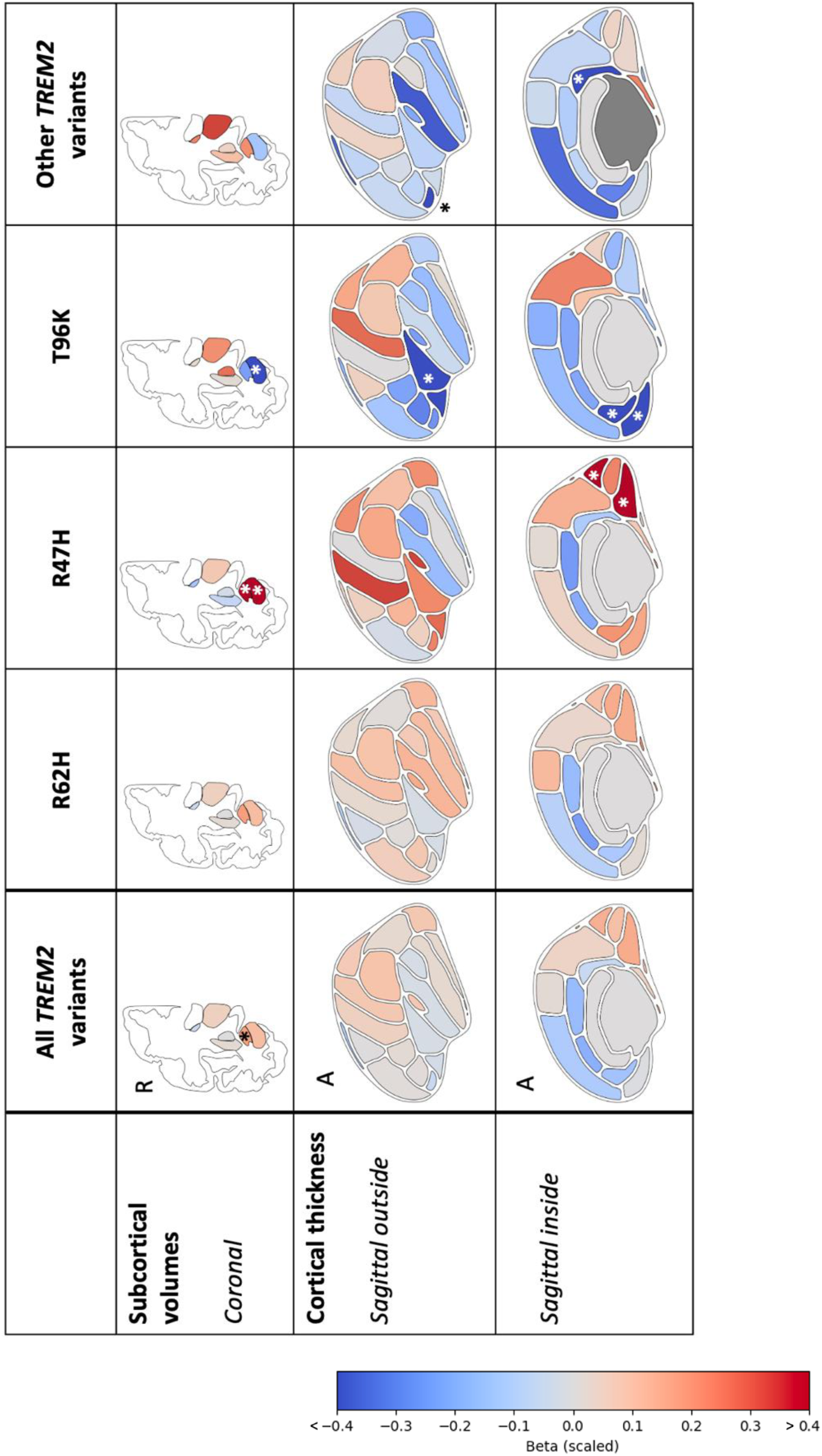
*TREM2* effect on cortical thickness and subcortical volumes measured on quantitative imaging analyses using MRI compared to non-carriers. Shown here are standardized betas adjusted for age, sex, and estimated intracranial volume. Other *TREM2* variants include: D87N, G58A, Q33X, C51Y, R47G, R31F, and A105V. Abbreviations: A = Anterior; R47H = p.Arg47His; R62H = p.Arg62His; p.T96K = Thr96Lys; R = Right; *TREM2* = Triggering Receptor Expressed on Myeloid Cells 2.

**Fig. 6:**
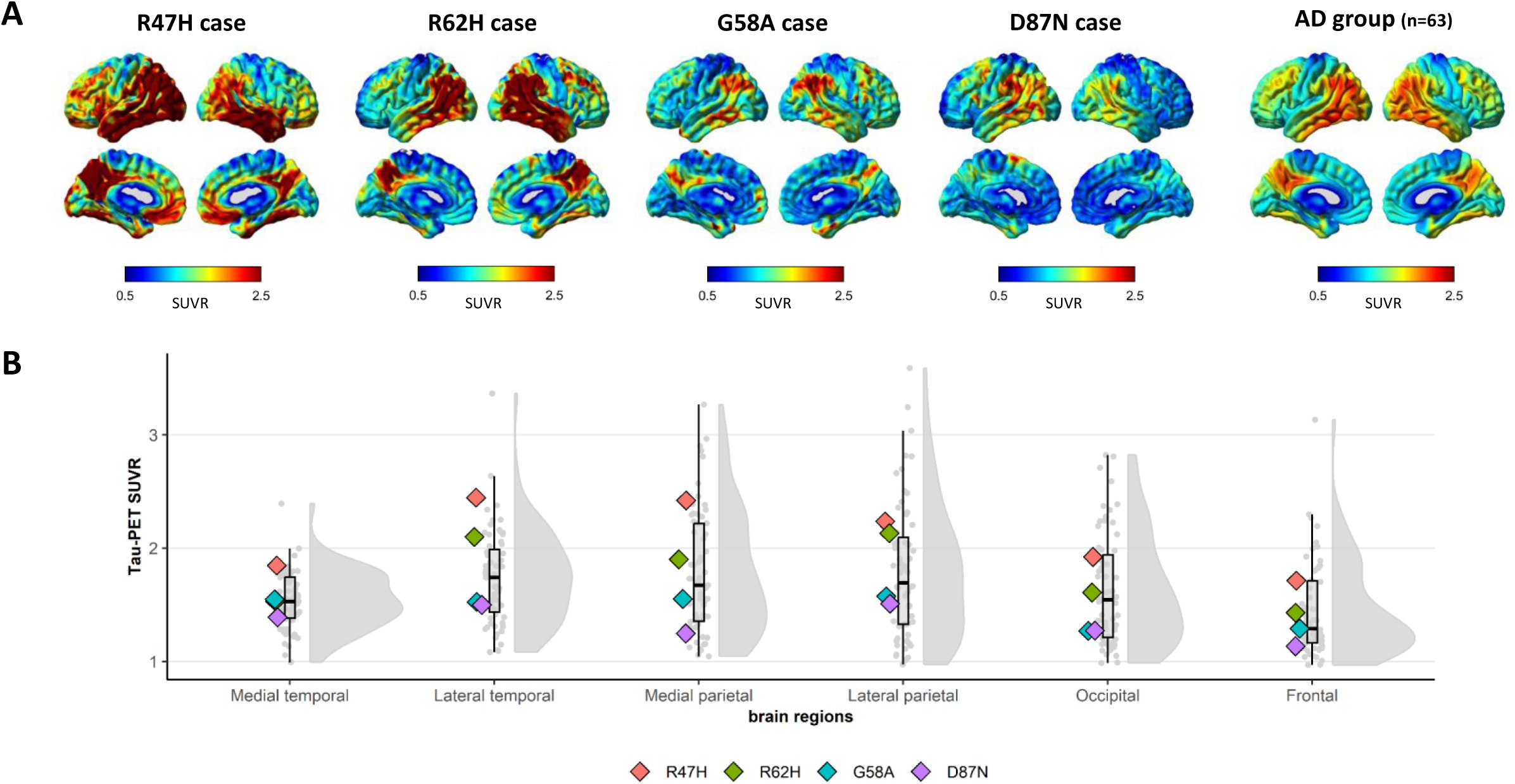
[^18^F] flortaucipir PET scans of *TREM2* variant carriers compared to early-onset AD non-carriers. Abbreviations: D87N = p.Asp87Asn; G58A = p.Gly58Ala ; R47H = p.Arg47His; R62H = p.Arg62His; *TREM2* = Triggering Receptor Expressed on Myeloid Cells 2.

### Exploratory analysis of specific *TREM2* variants

#### Effects of TREM2 R62H

Carriers of the R62H variant (*n*=66, 54% of *TREM2* carriers) showed less impaired scores in attention and speed compared to AD patients not carrying a *TREM2* variant (std*β* −0.28 ±0.14, *P*=0.042). R62H carriers did not show different CSF core AD biomarker levels. On MRI, R62H carriers had less white matter intensities, and less atrophy in the temporal pole compared to non-carriers (Fazekas std*β* - 0.27 ±0.13, *P*=3.5×10^−2^; std*β* 0.29 ±0.14, *P*=3.7×10^−2^), but no difference in other atrophy measures (i.e., MTA, PCA, 40 ROIs). R62H carriership did not associate with EEG abnormality. Longitudinally, the R62H variant did not show a significant effect on cognitive decline as measured by MMSE (*β* −0.54 ± 0.31, *P*=8.5×10^−2^), or on time between diagnosis and death (HR *P*=0.76).

#### Effects of TREM2 R47H

Carriers of the R47H variant (*n*=26, 21% of *TREM2* carriers) showed more impaired scores in language and global cognition compared to AD patients not carrying a *TREM2* variant (std*β* −0.38 ±0.17, *P*=2.7×10^−2^ and std*β* −0.56 ±0.16, *P*=4.4×10^−4^). R47H carriers had higher CSF-pTau181 and t-tau levels compared to non-carriers (std*β* 0.60 ±0.20, *P*=2.7×10^−3^ and std*β* 0.47 ±0.20, *P*=1.8×10^−2^). No effect was found in Aβ42 or NfL levels. On MRI, R47H carriers had less atrophy in the hippocampus and amygdala (std*β* 0.49 ±0.20, *P*=1.6×10^−2^ and std*β* 0.69 ±0.21, *P*=9.5×10^−4^), whereas the global temporal regions tended to show (non-significantly) more atrophy compared to non-carriers. The lingual and cuneus regions in the occipital lobe showed less cortical atrophy compared to non-carriers (std*β* 0.47 ±0.24, *P*=4.9×10^−2^ and std*β* 0.49 ±0.24, *P*=3.7×10^−2^). R47H carriership did not associate with MRI visual ratings or EEG visual scores. Longitudinally, R47H carriers showed a faster cognitive decline compared to non-carriers (−3.2 points decline per year of follow-up, β-difference −1.4 ±0.61, *P*=2.1×10^−2^, Fig. 4B). Moreover, R47H carriers were at increased risk of mortality (HR 1.60, 95% CI 1.0-2.5, *P*=3.4×10^−2^).

#### Effects of TREM2 T96K

Carriers of the T96K variant (*n*=16, 13% of *TREM2* carriers) had more impaired memory and language compared to AD patients not carrying a *TREM2* variant (std*β* −0.42 ±0.18, *P*=1.8×10^−2^ and std*β* −0.37 ±0.18, *P*=3.4×10^−2^). T96K carriership was associated with lower levels of CSF-Aβ42, pTau-181, and t-tau (std*β* −0.64 ±0.21, *P*=2.8×10^−3^, std*β* −0.50 ±0.21, *P*=1.9×10^−2^ and std*β* −0.45 ±0.21, *P*=3.5×10^−2^), but not with different NfL levels compared to non-carriers. On MRI, we observed that T96K carriers had better parietal cortical atrophy scores (std*β* −0.56 ±0.23, *P*=1.3×10^−2^). T96K carriers showed more hippocampal atrophy (std*β* −0.64 ±0.22, *P*=3.3×10^−3^), which tended to expand (non-significantly) into the temporal lobe. T96K carriership did not associated with EEG abnormality. Longitudinally, carriers of T96K showed faster cognitive decline (β-difference −1.75 ±0.74, *P*=1.8×10^−2^). The T96K variant did not show a significant effect on time between diagnosis and death (HR 0.66, 95% CI 0.3-1.5, *P*=3.1×10^−1^).

#### Effects of other TREM2 risk variants

Carriers of other *TREM2* variants (*n*=17, 14% of *TREM2* carriers; D87N, G58A, Q33X, C51Y, R47G, R31F, and A105V) were grouped due to small sample sizes. These carriers were not significantly different on neuropsychological domains, MRI visual ratings, or the CSF core AD biomarker levels than AD patients not carrying a *TREM2* variant. On structural MRI, carriers of other *TREM2* variants showed more atrophy in the frontal region (pars orbitalis: std*β* −0.59 ±0.27, *P*=3.3×10^−2^, frontal pole: std*β* −0.56 ±0.28, *P*=4.2×10^−2^) and posterior cingulate region than non-carriers (isthmus cingulate: std*β* −0.61 ±0.28, *P*=2.7×10^−2^). Carriers of other *TREM2* variants were not associated with EEG abnormality. Longitudinally, other *TREM2* variants did not show a significant effect on cognitive decline (β-difference 0.58 ±0.62, *P*=3.5×10^−1^), or on time between diagnosis and death (*P*=7.1×10^−1^).

### Sensitivity analysis

After removal of population outliers, familial relations, and adjusting the models for three principal components, the cohort consisted of *n*=103 *TREM2* variant carriers and *n*=1,341 non-carriers. In the main analysis, the *TREM2* effect of less impaired visuospatial functioning and faster cognitive decline remained, albeit non-significantly (std*β* 0.22 ±0.1, *P*=1.4×10^−1^, and β −0.52 ±0.3, *P*=2.5×10^−1^; Supplementary Table 6). In the exploratory analysis, the association of R62H (*n*=60 carriers) with less white matter intensities (std*β* −0.30 ±0.1, *P*=2.5×10^−2^) remained. The less impaired attention and speed, and less atrophy in the temporal pole were non-significant but in the same direction (std*β* 0.08 ±0.1; std*β* 0.25 ±0.1). The effects seen in R47H (*n*=26 carriers) remained, i.e., more impaired global cognition (std*β* −0.56 ±0.2, *P*=4.4×10^−4^), higher pTau-181 and t-tau levels (std*β* 0.60 ±0.2, *P*=2.6×10^−3^, std*β* 0.46 ±0.2, *P*=2.0×10^−2^), less atrophy in the hippocampus and amygdala, and less atrophy in the posterior lobe (std*β* 0.47 ±0.2, *P*=2.3×10^−2^, std*β* 0.70 ±0.2, *P*=1.0×10^−2^, cuneus: std*β* 0.48 ±0.2, *P*=4.1×10^−2^), faster cognitive decline (β −1.4 ±0.6, *P*=2.2×10^−2^), and less time between diagnosis and death (β 0.46 ±0.2, *P*=3.8×10^−2^). The effects seen in T96K could not be calculated as only three T96K carriers were of European ancestry. This was anticipated as T96K is more prevalent in individuals with an African genetic ancestry. The effects seen in other *TREM2* variants (*n*=16 carriers) with atrophy in the frontal region (pars orbitalis: std*β* −0.55 ±0.28, and frontal pole: std*β* - 0.54 ±0.28, albeit non-significantly) and posterior cingulate region (isthmus cingulate: std*β* −0.61 ±0.28) remained.

## DISCUSSION

This study gives an overview of *TREM2*-associated and variant-specific clinical measures in symptomatic Alzheimer’s disease (*n*=1,582 including 7.8% *TREM2* variant carriers). Our primary finding was that *TREM2* variant carriers do not show a clinically distinct profile at baseline measures compared to patients with AD who do not carry a *TREM2* variant, however they do show faster cognitive decline in follow-up. This was most obvious in R47H and T96K carriers who progress nearly twice as fast as non-carriers. The more pronounced cognitive decline in R47H carriers (*n*=26) was accompanied by a shorter time between diagnosis and death, more impaired global cognition, higher CSF-pTau181 and t-tau levels, but with relative sparing of the hippocampal volume, as was previously observed in post-mortem studies (17). The more pronounced cognitive decline in T96K carriers (*n*=16) was accompanied by more impaired language, lower levels of the core AD biomarkers CSF-Aβ42, pTau-181 and t-tau, and more hippocampal and temporal atrophy. In summary, *TREM2* variants carriers, especially R47H and T96K, seemed to have a more aggressive form of AD and the underlying biological mechanism of faster progression could differ between *TREM2* variants. This knowledge could help us understand the effects of the *TREM2* gene, enrich clinical trials for fast progressors, and inform the development of future TREM2 therapies.

### *TREM2* variant carriers as fast progressors

Our findings support that *TREM2* variation is involved in processes relevant for cognitive decline. Already in the discovery of *TREM2*, R47H showed worse cognition as a function of age than non-carriers (1), although this study did not differentiate between progression after a diagnosis of AD dementia. Another study by Kim et al. (2022) also reported faster cognitive decline in *TREM2* variant carriers (*n*=12 of whom *n*=8 R47H carriers) compared to AD non-carriers (17). R47H and T96K carriers showed twice as fast cognitive decline than non-carriers. Hence, this suggests that specifically R47H and T96K carriers, as fast progressors, are interesting candidates for enrichment in *TREM2*-targeting clinical trials for AD. As T96K carriership is common in African ancestry (12.5% of the African (American) population) (64), targeting this subgroup could offer valuable insights within diverse populations affected by AD. To conclude, the observed *TREM2* effects should be considered in studies of disease progression such as clinical trials. This is particularly important when treatment groups are enriched with individuals of African ancestry, as the faster progression induced by the T96K variant, present in over 12% of this population, may even mask a treatment effect.

### *TREM2* carriers present as typical AD

The pattern and severity of cognitive impairments can vary among individuals with AD (65). Our study did not identify any distinct patterns of AD. However, we did observe less impaired visuospatial functioning among carriers of a *TREM2* variant compared to non-carriers. One possible explanation for this discrepancy could be that visuospatial difficulties manifest later in the disease progression of *TREM2* variant carriers compared to non-carriers. The mechanism underlying the observed differences in visuospatial functioning among *TREM2* variant carriers with AD remains unclear and warrants further research, especially considering the complex interplay of brain regions and networks involved in visuospatial functioning (66) and the borderline significance of the FDR-corrected p-value.

### *TREM2* R47H effect on tau

R47H carriers in our cohort showed higher CSF-pTau181 and t-tau levels compared to other variants in *TREM2*. This is in line with a GWAS study on CSF, which reported a strong association between AD patients carrying this variant and higher levels of CSF-pTau181 and tau compared to AD non-carriers (67). In terms of brain atrophy, we found preserved volumes of hippocampus and amygdala and less occipital atrophy than non-carriers. Pathology findings on *TREM2* variant carriers align with our R47H findings and reported an overall higher tau burden than AD non-carriers, no altered Aβ burden, and a significantly lower tau burden in hippocampal regions (17,18). Together, this could suggest faster tau accumulation in the brain of R47H carriers than non-carriers (68), as well as a stronger down-stream effect of amyloid.

As tau is a predictor of disease progression as shown in tau-PET studies (69,70), this makes TREM2 an interesting target for disease-modifying therapies to slow progression of disease by enhancing TREM2 activation (15). However, even though R47H carriers showed higher CSF-pTau181 and t-tau levels, T96K carriers showed lower pTau181 and t-tau levels suggesting another mechanism of tau processing. As *TREM2* variants impact tau, this genetic factor could modify the effect of disease-modifying therapies. Hence, *TREM2* variants could be considered when evaluating the effect of such therapies.

### Strengths and limitations

The main strength of the study is the use of a large monocentre clinical dataset of *TREM2* variant carriers with available data from all clinical measures. This dataset facilitated precise estimations of *TREM2* effects on multilayered phenotypes and disease progression. The large sample also enabled exploratory analyses of the separate *TREM2* variants. In addition, the consistency of the diagnostic trajectory across all patients from 2000 to 2023 prevents ascertainment bias (31). Moreover, the strict inclusion criteria, limited to amyloid-confirmed AD patients, and the thorough identification of non-carriers through WES of the *TREM2* gene increased the homogeneity of the data and likely the reliability of results. Moving forward, the results of the exploratory analysis should be replicated in other cohorts including more diverse populations. Specifically, the T96K effect on cognitive decline should be replicated by comparing carriers and non-carriers of African ancestry.

Our findings suggest that specific *TREM2* variants can influence the disease phenotype and progression, highlighting the importance of genetic factors in AD. This knowledge can enhance the understanding of the molecular mechanisms underlying AD and support the development of targeted therapies. Additionally, the observed *TREM2* variant-specific effects could be considered as a factor to be included in inclusion criteria to improve clinical trial design and evaluation of the effect of such therapies, potentially leading to more personalized treatment approaches.

## DATA AVAILABILITY

Data is provided within the manuscript or supplementary information files. The dataset used and/or the analyses performed can be provided upon reasonable request from data manager of the ADC (W.F.).

## ACKNOWLEDGEMENTS

We thank all study participants and all personnel involved in data collection for the contributing studies. S.L. was funded for this study by NWO (#733050512, PROMO-GENODE: a PROspective study of MOnoGEnic causes Of Dementia) a substantial donation by Edwin Bouw Fonds, Dioraphte and YOD-INCLUDED (ZonMW project no. 10510032120002), and S.L. is part of the Dutch Dementia Research Programme. S.L. further received funding for the GeneMINDS consortium, which is powered by Health∼Holland, Top Sector Life Sciences & Health. Research of Alzheimer Center Amsterdam is part of the neurodegeneration research program of Amsterdam Neuroscience. Alzheimer Center Amsterdam is supported by Stichting Alzheimer Nederland and Stichting Steun Alzheimercentrum Amsterdam. The chair W.F. is supported by the Pasman stichting. W.F., S.L., H.H., M.H. are recipients of ABOARD, which is a public-private partnership receiving funding from ZonMW (#73305095007) and Health∼Holland, Topsector Life Sciences & Health (PPP-allowance; #LSHM20106). More than <u>30 partners</u> participate in ABOARD. ABOARD also receives funding from de Hersenstichting, Edwin Bouw Fonds and Gieskes-Strijbisfonds. Array genotyping was performed in the context of EADB (European Alzheimer DNA biobank) funded by the JPco-fuND FP-829-029 (ZonMW projectnumber 733051061). V.V. is supported by JPND-funded E-DADS project (ZonMW project #733051106). The work in this manuscript was carried out on the Snellius supercomputer, which is embedded in the Dutch national e-infrastructure with the support of SURF Cooperative. Computing hours were granted in 2016, 2017, 2018 and 2019 to H.H. by the Dutch Research Council (project name: ‘100plus’; project numbers 15318 and 17232). This work also used the Dutch national e-infrastructure with the support of the SURF Cooperative using grant no. EINF-2044 and EINF-5353, granted to V.V. F.B. is supported by the NIHR biomedical research centre at UCLH.

## AUTHOR INFORMATION

These authors jointly supervised this work: Lisa Vermunt, Sven van der Lee.

### Author’s contributions

J.D., L.V., and S.v.d.L. designed the study, had full access to the raw data, carried out the final statistical analyses, wrote the manuscript, and had the final responsibility to submit for publication. All authors contributed either demographic, clinical, genetic, biomarker, or neuroimaging data. All authors contributed to the interpretation of the results, critically reviewed the manuscript, and approved the final manuscript.

## ETHICS DECLARATIONS

### Ethics approval and consent to participate

The study was approved by the Medical Ethical Committee of Amsterdam UMC, location VUmc. All patients provided written informed consent for their clinical data to be used for research purposes. Consent was obtained according to the Declaration of Helsinki.

## REFERENCES

1. Jonsson T, Stefansson H, Steinberg S, Jonsdottir I, Jonsson P V, Snaedal J, et al. Variant of TREM2 Associated with the Risk of Alzheimer’s Disease. N Engl J Med. 2013 Jan 10;368(2):107–23.

2. Guerreiro R, Wojtas A, Bras J, Carrasquillo M, Rogaeva E, Majounie E, et al. TREM2 Variants in Alzheimer’s Disease. N Engl J Med. 2013 Jan 1;368(2):117.

3. Holstege H, Hulsman M, Charbonnier C, Grenier-Boley B, Quenez O, Grozeva D, et al. Exome sequencing identifies rare damaging variants in ATP8B4 and ABCA1 as risk factors for Alzheimer’s disease. Nat Genet. 2022 Nov 21.

4. Sherva R, Zhang R, Sahelijo N, Jun G, Anglin T, Chanfreau C, et al. African ancestry GWAS of dementia in a large military cohort identifies significant risk loci. Mol Psychiatry. 2022 Dec 22;28:1293–302.

5. Bis JC, Jian X, Kunkle BW, Chen Y, Hamilton-Nelson KL, Bush WS, et al. Whole exome sequencing study identifies novel rare and common Alzheimer’s-Associated variants involved in immune response and transcriptional regulation. Mol Psychiatry [Internet]. 2020 [cited 2024 Apr 25];24:1859–75.

6. ≈ R, van der Lee SJ, Naj AC, Bellenguez C, Badarinarayan N, Jakobsdottir J, et al. Rare coding variants in PLCG2, ABI3, and TREM2 implicate microglial-mediated innate immunity in Alzheimer’s disease. Nature Publishing Group. 2017 Sep;49(9):1373–87.

7. Gratuze M, Leyns CEG, Holtzman DM. New insights into the role of TREM2 in Alzheimer’s disease. Vol. 13, Molecular Neurodegeneration. BioMed Central Ltd.; 2018.

8. Ulland TK, Colonna M. TREM2 — a key player in microglial biology and Alzheimer disease. Nat Rev Neurol. 2018 Nov;14:667–75.

9. Scheltens P, De Strooper B, Kivipelto M, Holstege H, Chételat G, Teunissen CE, et al. Alzheimer’s disease. Lancet. 2021 Apr 24;397(10284):1577–90.

10. Kinney JW, Bemiller SM, Murtishaw AS, Leisgang AM, Salazar AM, Lamb BT. Inflammation as a central mechanism in Alzheimer’s disease. Alzheimer’s and Dementia: Translational Research and Clinical Interventions. 2018 Jan 1;4:575–90.

11. Van Bokhoven P, De Wilde A, Vermunt L, Leferink PS, Heetveld S, Cummings J, et al. The Alzheimer’s disease drug development landscape. Alzheimers Res Ther. 2021;13.

12. Leng F, Edison P. Neuroinflammation and microglial activation in Alzheimer disease: where do we go from here? Nat Rev Neurol. 2021 Mar;17:157–72.

13. Cisbani G, Rivest S. Targeting innate immunity to protect and cure Alzheimer’s disease: opportunities and pitfalls. Mol Psychiatry. 2021;26:5504–15.

14. Cummings J, Zhou Y, Lee G, Zhong K, Fonseca J, Cheng F. Alzheimer’s disease drug development pipeline: 2023. Alzheimer’s & Dementia: Translational Research & Clinical Interventions. 2023 May 25.

15. Lengerich B, Zhan L, Xia D, Chan D, Joy D, Park JI, et al. A TREM2-activating antibody with a blood-brain barrier transport vehicle enhances microglial metabolism in Alzheimer’s disease models. Nat Neurosci. 2023;26:416–29.

16. Li RY, Qin Q, Yang HC, Wang YY, Mi YX, Yin YS, et al. TREM2 in the pathogenesis of AD: a lipid metabolism regulator and potential metabolic therapeutic target. Vol. 17, Molecular Neurodegeneration. BioMed Central Ltd; 2022.

17. Kim B, Suh E, Nguyen AT, Prokop S, Mikytuck B, Olatunji OA, et al. TREM2 risk variants are associated with atypical Alzheimer’s disease. Acta Neuropathol. 2022;144:1085–102.

18. Prokop S, Miller KR, Labra SR, Pitkin RM, Hoxha K, Narasimhan S, et al. Impact of TREM2 risk variants on brain region-specific immune activation and plaque microenvironment in Alzheimer’s disease patient brain samples. Acta Neuropathol. 2019;138:613–30.

19. Luis E, Ortega-Cubero S, Lamet I, Razquin C, Cruchaga C, Benitez B, et al. Frontobasal gray matter loss is associated with the TREM2 p.R47H variant. Neurobiological Aging. 2014;35(12):2681–90.

20. Devi G, Scheltens P. Heterogeneity of Alzheimer’s disease: Consequence for drug trials? Vol. 10, Alzheimer’s Research and Therapy. BioMed Central Ltd.; 2018.

21. Jay TR, Von Saucken VE, Landreth GE. TREM2 in Neurodegenerative Diseases. Mol Neurodegener. 2017 Aug 2;12.

22. Fancy N, Willumsen N, Tsartsalis S, Khozoie C, Mcgarry A, Muirhead RC, et al. Mechanisms contributing to differential genetic risks for TREM2 R47H and R62H variants in Alzheimer’s Disease. 2022.

23. Kober DL, Alexander-Brett JM, Karch CM, Cruchaga C, Colonna M, Holtzman MJ, et al. Neurodegenerative disease mutations in TREM2 reveal a functional surface and distinct loss-of-function mechanisms. Biophysics and Structural Biology. 2016;5:1–24.

24. Song W, Hooli B, Mullin K, Chih Jin S, Cella M, Ulland TK, et al. Alzheimer’s disease-associated TREM2 variants exhibit either decreased or increased ligand-dependent activation. Alzheimer’s Dementia. 2017;13(4):381–7.

25. Del-Aguila JL, Benitez BA, Li Z, Dube U, Mihindukulasuriya KA, Budde JP, et al. TREM2 brain transcript-specific studies in AD and TREM2 mutation carriers. Mol Neurodegener. 2019;14(18).

26. Tijms BM, Vromen EM, Mjaavatten O, Holstege H, Reus LM, van der Lee S, et al. Large-scale cerebrospinal fluid proteomic analysis in Alzheimer’s disease patients reveals five molecular subtypes with distinct genetic risk profiles. Nat Aging. 2024;4:33–47.

27. Slattery CF, Beck JA, Harper L, Adamson G, Abdi Z, Uphill J, et al. R47H TREM2 variant increases risk of typical early-onset Alzheimer’s disease but not of prion or frontotemporal dementia. Alzheimer’s & Dementia. 2014 Nov 1;10(6):602–8.

28. Lill CM, Rengmark A, Pihlstrøm L, Fogh I, Shatunov A, Sleiman PM, et al. The role of TREM2 R47H as a risk factor for Alzheimer’s disease, frontotemporal lobar degeneration, amyotrophic lateral sclerosis, and Parkinson’s disease. Alzheimer’s and Dementia. 2015 Dec 1;11(12):1407–16.

29. Slattery C, Beck J, Harper L, Adamson G, Abdi Z, Campbell T, et al. R47H TREM2 variant increases risk of typical early-onset Alzheimer’s disease but not of prion or frontotemporal dementia. Alzheimers Dementia. 2014 Nov;10(6):602–8.

30. Dokholyan N V., Mohs RC, Bateman RJ. Challenges and progress in research, diagnostics, and therapeutics in Alzheimer’s disease and related dementias. Alzheimer’s and Dementia: Translational Research and Clinical Interventions. 2022;8:1–5.

31. Van Der Flier WM, Scheltens P. Amsterdam Dementia Cohort: Performing Research to Optimize Care. Journal of Alzheimer’s Disease. 2018 Jan 1;62(3):1091–111.

32. Bellenguez C, Küçükali F, Jansen IE, Kleineidam L, Moreno-Grau S, Amin N, et al. New insights into the genetic etiology of Alzheimer’s disease and related dementias. Nat Genet. 2022;54:412–36.

33. Das S, Forer L, Schönherr S, Sidore C, Locke AE, Kwong A, et al. Next-generation genotype imputation service and methods. Nat Genet. 2016 Oct;48(10).

34. Taliun D, Harris DN, Kessler MD, Carlson J, Szpiech ZA, Torres R, et al. Sequencing of 53,831 diverse genomes from the NHLBI TOPMed Program. Nature. 2021.

35. Tesi N, Lee SJ van der, Hulsman M, Jansen IE, Stringa N, Schoor N Van, et al. Centenarian controls increase variant effect sizes by an average twofold in an extreme case-extreme control analysis of Alzheimer’s disease. European Journal of Human Genetics. 2019;27:244–53.

36. Wang L, Nykänen NP, Western D, Gorijala P, Timsina J, Li F, et al. Proteo-genomics of soluble TREM2 in cerebrospinal fluid provides novel insights and identifies novel modulators for Alzheimer’s disease. Mol Neurodegener. 2024;19:1.

37. Ioannidis NM, Rothstein JH, Pejaver V, Middha S, Mcdonnell SK, Baheti S, et al. An Ensemble Method for Predicting the Pathogenicity of Rare Missense Variants. The American Journal of Human Genetics. 2016;99:877–85.

38. Folstein MF, Folstein SE, McHugh PR. “Mini-mental state”: A practical method for grading the cognitive state of patients for the clinician. J Psychiatr Res. 1975 Nov 1;12(3):189–98.

39. Dubbelman MA, Hendriksen HMA, Harrison JE, Vijverberg EGB, Prins ND, Kroeze LA, et al. Cognitive and Functional Change over Time in Cognitively Healthy Individuals According to Alzheimer Disease Biomarker-Defined Subgroups. Neurology. 2024 Jan 23;102(2).

40. Tijms BM, Willemse EAJ, Zwan MD, Mulder SD, Visser PJ, Van Berckel BNM, et al. Unbiased Approach to Counteract Upward Drift in Cerebrospinal Fluid Amyloid-B 1– 42. Clin Chem. 2018;64(3):576–85.

41. Duits FH, Teunissen CE, Bouwman FH, Visser PJ, Mattsson N, Zetterberg H, et al. The cerebrospinal fluid “alzheimer profile”: Easily said, but what does it mean? Alzheimer’s and Dementia. 2014 Nov 1;10(6):713–723.e2.

42. Schindler SE, Gray JD, Gordon BA, Xiong C, Batrla-Utermann R, Quan M, et al. Cerebrospinal fluid biomarkers measured by Elecsys assays compared to amyloid imaging. Alzheimer’s and Dementia. 2018 Nov 1;14(11):1460–9.

43. Willemse EAJ, van Maurik IS, Tijms BM, Bouwman FH, Franke A, Hubeek I, et al. Diagnostic performance of Elecsys immunoassays for cerebrospinal fluid Alzheimer’s disease biomarkers in a nonacademic, multicenter memory clinic cohort: The ABIDE project. Alzheimer’s and Dementia: Diagnosis, Assessment and Disease Monitoring. 2018 Jan 1;10:563–72.

44. Vermunt L, Otte M, Verberk IMW, Killestein J, Lemstra AW, van der Flier WM, et al. Age- and disease-specific reference values for neurofilament light presented in an online interactive support interface. Ann Clin Transl Neurol. 2022 Nov 1;9(11):1832–7.

45. Scheltens P, Leys D, Barkhof F, Huglo D, Weinstein HC, Vermersch P, et al. Atrophy of medial temporal lobes on MRI in “probable” Alzheimer’s disease and normal ageing: diagnostic value and neuropsychological correlates. J Neurol Neurosurg Psychiatry. 1992;55:967–72.

46. Koedam ELGE, Lehmann M, Van Der Flier WM, Scheltens P, Pijnenburg YAL, Fox N, et al. Visual assessment of posterior atrophy development of a MRI rating scale. Eur Radiology. 2011 Mar 18;21:2618–25.

47. Fazekas F, Chawluk JB, Alavi A, Hurtig HI, Zimmerman RA. MR signal abnormalities at 1.5 T in Alzheimer’s dementia and normal aging. American Journal of Roentgenology. 1987;149(2):351–6.

48. Briels CT, Stam CJ, Scheltens P, Gouw AA. The predictive value of normal EEGs in dementia due to Alzheimer’s disease. Ann Clin Transl Neurol. 2021 May 1;8(5):1038–48.

49. Liedorp M, Van Der Flier WM, Hoogervorst ELJ, Scheltens P, Stam CJ. Associations between Patterns of EEG Abnormalities and Diagnosis in a Large Memory Clinic Cohort. Original Research Article Dement Geriatr Cogn Disord. 2009;27:18–23.

50. Fischl B. FreeSurfer. Neuroimage. 2012 Aug 15;62(2):774–81.

51. Archetti D, Venkatraghavan V, Weiss B, Bourgeat P, Auer T, Vidnyánszky Z, et al. A machine-learning model to harmonize brain volumetric data for quantitative neuro-radiological. medRxiv. 2024 Feb 3.

52. Bocancea DI, den Braber A, Jiang C, Coomans EM, van Unnik AAJM, van Veen JML, et al. Automated FreeSurfer segmentation and visual quality control in 10,000 MRI scans from a large memory clinic cohort. Alzheimer’s & Dementia. 2023 Dec;19(S16).

53. Fortin JP, Cullen N, Sheline YI, Taylor WD, Aselcioglu I, Cook PA, et al. Harmonization of cortical thickness measurements across scanners and sites. Neuroimage. 2018 Feb 15;167:104–20.

54. Visser D, Wolters EE, J Verfaillie SC, Coomans EM, Timmers T, Tuncel H, et al. Tau pathology and relative cerebral blood flow are independently associated with cognition in Alzheimer’s disease. Eur J Nucl Med Mol Imaging. 2020;47:3165–75.

55. Wolters EE, van de Beek M, Ossenkoppele R, Verfaillie SCJ, Coomans EM, Timmers T, et al. Tau pathology, relative cerebral flow and cognition in dementia with Lewy bodies. Alzheimer’s & Dementia. 2020 Dec;16(S1).

56. Golla SS V, Timmers T, Ossenkoppele R, Groot C, Verfaillie S, Scheltens P, et al. Quantification of Tau Load Using [18 F]AV1451 PET. Mol Imaging Biol. 2017;19:963–71.

57. Coomans EM, de Koning LA, Rikken RM, Verfaillie SC, Visser D, den Braber A, et al. Performance of a [^18^F] Flortaucipir PET Visual Read Method Across the Alzheimer Disease Continuum and in Dementia With Lewy Bodies. 2023;101:1850–62.

58. Singleton E, Hansson O, Pijnenburg YAL, Joie R La, Mantyh WG, Tideman P, et al. Heterogeneous distribution of tau pathology in the behavioural variant of Alzheimer’s disease. J Neurol Neurosurg Psychiatry. 2021;1–9.

59. R: A language and environment for statistical computing. 2023.

60. Python. 2024.

61. F. Verhage. Intelligentie en leeftijd; onderzoek bij Nederlanders van twaalf tot zevenenzeventig jaar. 1964.

62. European Commission. International Standard Classification of Education (ISCED). 2023.

63. The 1000 Genomes Project Consortium. A global reference for human genetic variation. Nature. 2015 Oct 1;526.

64. gnomAD v4.1.0. Genome Aggregation Database. 2024.

65. Qiu Y, Jacobs DM, Messer K, Salmon DP, Feldman HH, Alzheimer’s Disease SM, et al. Cognitive heterogeneity in probable Alzheimer disease. Neurology. 2019;93:778–90.

66. Mandal PK, Joshi J, Saharan S. Visuospatial perception: An emerging biomarker for Alzheimer’s disease. Vol. 31, Journal of Alzheimer’s Disease. IOS Press; 2012. p. 117–35.

67. Cruchaga C, Kauwe JSK, Harari O, Jin SC, Cai Y, Karch CM, et al. GWAS of cerebrospinal fluid tau levels identifies novel risk variants for Alzheimer’s disease. Neuron. 2013 Apr 4;78(2):256–68.

68. Sanchez-Mejias E, Navarro V, Jimenez S, Sanchez-Mico M, Sanchez-Varo R, Nuñez-Diaz C, et al. Soluble phospho-tau from Alzheimer’s disease hippocampus drives microglial degeneration. Acta Neuropathol. 2016 Dec 1;132(6):897–916.

69. Ossenkoppele R, Pichet Binette A, Groot C, Smith R, Strandberg O, Palmqvist S, et al. Amyloid and tau PET-positive cognitively unimpaired individuals are at high risk for future cognitive decline. Nat Med. 2022 Nov 1;28(11):2381–7.

70. Stephanie J. B. Vos, Frans Verhey, Lutz Frölich, Johannes Kornhuber, Jens Wiltfang, Wolfgang Maier, et al. New criteria for Alzheimer’s disease: Which, when and why? Brain. 2015 May 1;138(5):1134–7.

