## Supplementary Material for "*TREM2* Risk Variants with Alzheimer’s Disease Differ in Rate of Cognitive Decline"

**EXTENDED DATA**

**Extended Data Fig. 1 *TREM2* effect on cognitive decline stratified by *TREM2* mutation carriers compared to non-carriers.**

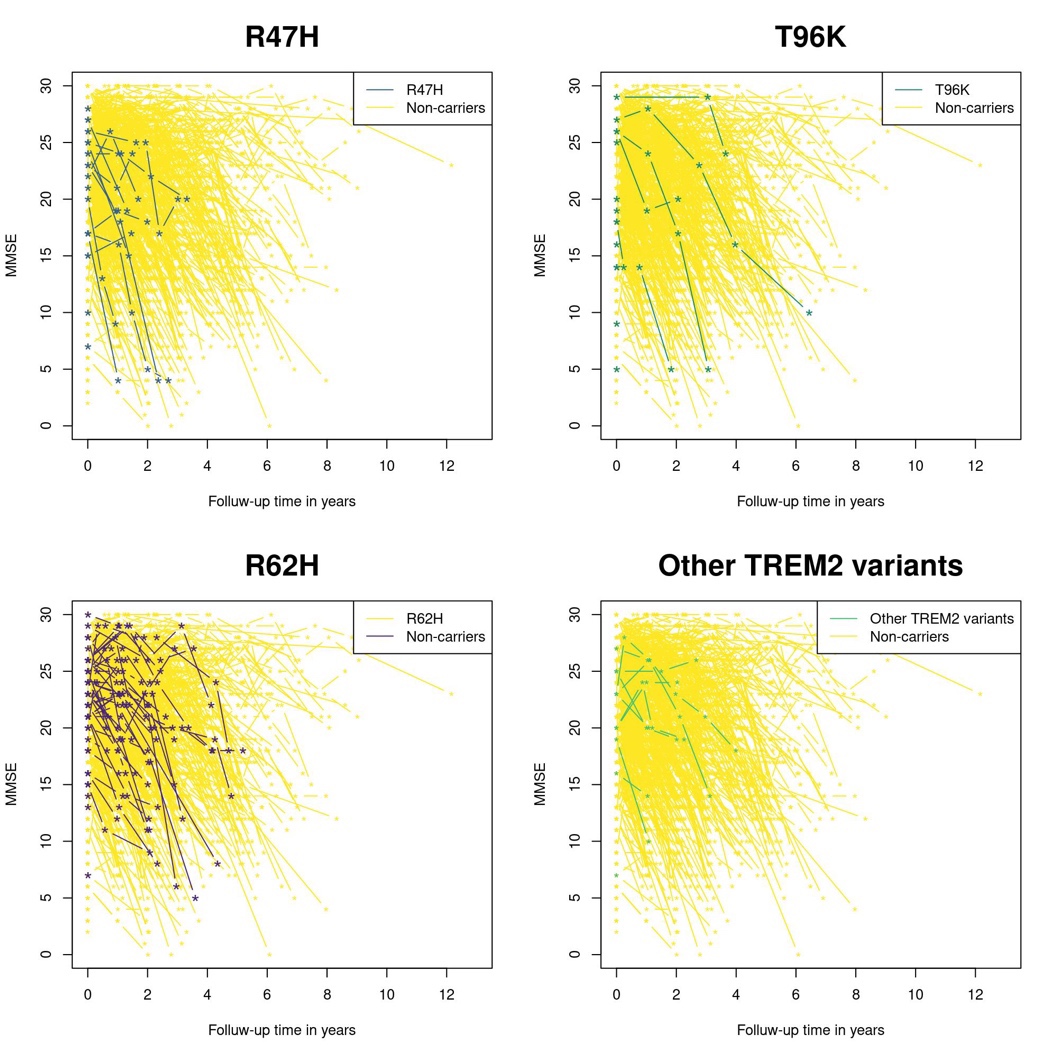

Shown here are MMSE values per individual over time, unadjusted.

Other *TREM2* variants include: D87N, G58A, Q33X, C51Y, R47G, R31F, and A105V.

Abbrev.: MMSE = Mini-Mental State Examination; R47H = p.Arg47His; R62H = p.Arg62His; T96K = p.Thr96Lys; *TREM2* = Triggering Receptor Expressed on Myeloid Cells 2.

**SUPPLEMENTARY MATERIAL**

**Supplementary Fig. 1: *TREM2* effect on time between diagnosis of symptomatic AD and death compared to non-carriers.**

(a)
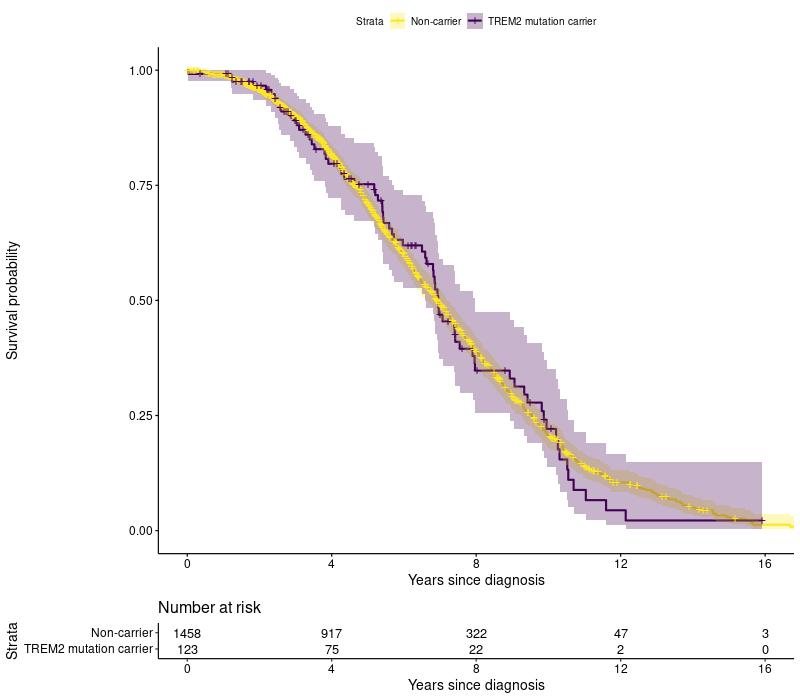

(b)
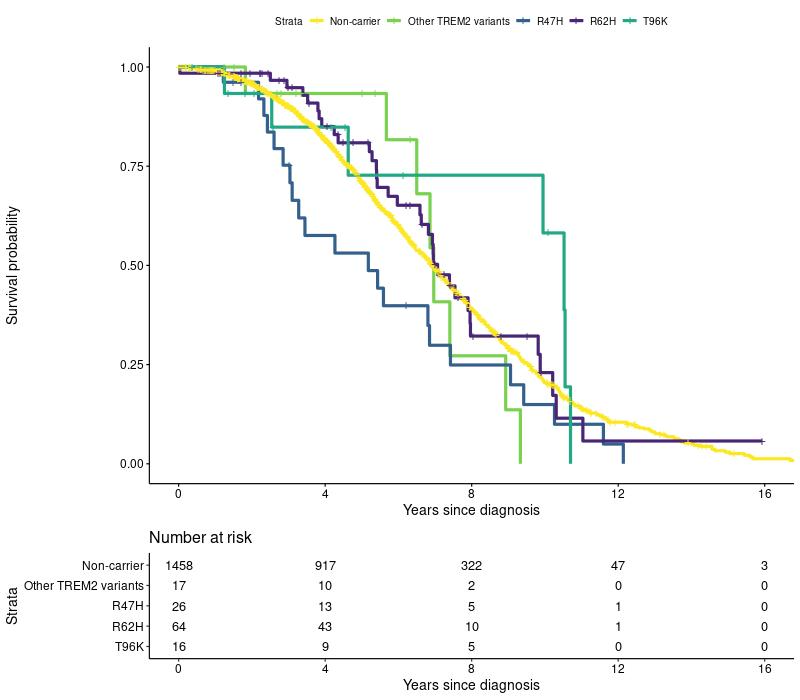

Shown here are unadjusted Kaplan Meijer curves of the time between diagnosis and death.

(a) *TREM2* mutation carriers vs. non-carriers. (b) *TREM2*-mutation carriers stratified by mutation vs. non-carriers.

Other *TREM2* variants include: D87N, G58A, Q33X, C51Y, R47G, R31F, and A105V.

Abbreviations: R47H = p.Arg47His; R62H = p.Arg62His; T96K = p.Thr96Lys; *TREM2* = Triggering Receptor Expressed on Myeloid Cells 2.

**Supplementary Table 1: Cohort characteristics per endophenotype.**

|  | **Main analysis** | |  |  |  |  |  |  | **Exploratory analysis** | |
| --- | --- | --- | --- | --- | --- | --- | --- | --- | --- | --- |
|  | Total | Neuropsych.  Domains | MRI clinical  ratings | CSF | CSF-NfL | EEG | Time Diagnosis  to Death | MMSE  over Time | MRI structural | Tau-PET |
| Total | 1582 (100) | 1519 (96) | 1210 (76) | 1522 (96) | 164 (10) | 1304 (82) | 1582 (100) | 1564 (99) | 1069 (68) | 67 (4) |
| Female | 824 (52) | 789 (52) | 637 (53) | 791 (52) | 94 (57) | 679 (52) | 824 (52) | 814 (52) | 558 (52) | 33 (49) |
| Age at diagnosis, *mean ±SD* | 64.4 ±7 | 64.4 ±7 | 64.8 ±7 | 64.4 ±7 | 63.4 ±7 | 64.4 ±7.1 | 64.4 ±7 | 64.4 ±7 | 64.6 ±7 | 63 ±7 |
| Education, *median (IQR)* | 1 (0-2) | 1 (0-2) | 1 (0-2) | 1 (0-2) | 1 (0-2) | 1 (0-2) | 1 (0-2) | 1 (0-2) | 1 (0-2) | 2 (1-2) |
| Positive family history^a^ | 691 (44) | 670 (44) | 524 (43) | 660 (43) | 78 (48) | 565 (43) | 691 (44) | 688 (44) | 466 (44) | 35 (52) |
| ApoE-ε4 carrier | 1076 (68) | 1044 (69) | 842 (70) | 1038 (68) | 115 (70) | 899 (69) | 1076 (68) | 1064 (68) | 743 (70) | 46 (69) |
| Died | 949 (60) | 894 (59) | 747 (62) | 924 (61) | 102 (62) | 811 (62) | 949 (60) | 934 (60) | 657 (61) | 35 (52) |
| Age at death | 71 ±8 | 71.1 ±8 | 71.5 ±8 | 71.1 ±7.7 | 69.2 ±8 | 71.2 ±8 | 71 ±8 | 71 ±8 | 71.2 ±8 | 69.4 ±7 |
| *TREM2* mutation carriers | 123 (8) | 118 (8) | 99 (8) | 117 (8) | 10 (6) | 105 (8) | 123 (8) | 120 (8) | 95 (9) | 4 (6) |
| R62H | 66 (54) | 64 (54) | 54 (55) | 61 (52) | 4 (40) | 57 (54) | 66 (54) | 66 (55) | 53 (56) | 1 (25) |
| R47H | 26 (21) | 25 (21) | 18 (18) | 26 (22) | 3 (30) | 23 (22) | 26 (21) | 24 (20) | 18 (19) | 1 (25) |
| T96K | 16 (13) | 14 (12) | 15 (15) | 16 (14) | 2 (20) | 12 (11) | 16 (13) | 15 (12) | 11 (12) | 0 (0) |
| Other variants* | 17 (14) | 17 (14) | 13 (13) | 16 (14) | 1 (10) | 15 (14) | 17 (14) | 17 (14) | 14 (15) | 2 (50) |

Table shows *n* (%) unless otherwise specified.

*Other *TREM2* variants include: D87N, G58A, Q33X, C51Y, R47G, R31F, and A105V.

a) Positive family history = affected first degree relative.

Abbreviations: AD = Alzheimer’s Disease; ApoE = Apolipoprotein E; CSF = cerebrospinal fluid; Diag. = Diagnosis; EEG = Electroencephalography; IQR = Interquartile Range; MMSE = Mini-Mental State Examination; MRI = Magnetic Resonance Imaging; n = number; NfL = Neurofilament Light; Neuropsych. = Neuropsychological; PET = Positron Emission Tomography; R47H = p.Arg47His; R62H = p.Arg62His; T96K = p.Thr96Lys; SD = Standard Deviation; *TREM2* = Triggering Receptor Expressed on Myeloid Cells 2.

**Supplementary Table 2:** Effect of *TREM2* variants on clinical measures in AD comparing *TREM2* mutation carriers vs. non-carriers, scaled.

|  | **Main analysis** | | **Exploratory analysis** | |  |  |  |  |  | |
| --- | --- | --- | --- | --- | --- | --- | --- | --- | --- | --- |
|  | All *TREM2* variants combined | | R62H |  | R47H |  | T96K |  | Other *TREM2* variants | |
|  | stdβ ± SE | p-value^✦^ | stdβ ± SE | p-value | stdβ ± SE | p-value | stdβ ± SE | p-value | stdβ ± SE | p-value |
| MMSE at baseline | -0.00 ±0.08 | 0.962 | 0.07 ±0.11 | 0.544 | -0.11 ±0.18 | 0.54 | -0.19 ±0.20 | 0.339 | 0.08 ±0.22 | 0.724 |
| Neuropsychological domains | |  |  |  |  |  |  |  |  |  |
| Memory | -0.14 ±0.07 | 0.247 | -0.10 ±0.10 | 0.327 | -0.27 ±0.16 | 0.097 | -0.42 ±0.18 | 0.018* | 0.13 ±0.19 | 0.487 |
| Executive functioning | 0.07 ±0.16 | 0.816 | 0.21 ±0.21 | 0.316 | -0.69 ±0.35 | 0.053 | -0.36 ±0.37 | 0.331 | 0.63 ±0.39 | 0.110 |
| Attention and speed | 0.12 ±0.10 | 0.667 | 0.28 ±0.14 | 0.042* | -0.40 ±0.22 | 0.072 | 0.18 ±0.25 | 0.463 | 0.07 ±0.25 | 0.774 |
| Language | -0.03 ±0.08 | 0.816 | 0.18 ±0.10 | 0.075 | -0.38 ±0.17 | 0.027* | -0.37 ±0.18 | 0.034* | -0.00 ±0.19 | 0.996 |
| Visuospatial functioning | 0.21 ±0.08 | 0.094^✦^ | 0.16 ±0.10 | 0.12 | 0.24 ±0.17 | 0.174 | 0.24 ±0.18 | 0.197 | 0.29 ±0.19 | 0.123 |
| Global cognition | -0.04 ±0.07 | 0.816 | 0.02 ±0.08 | 0.824 | -0.56 ±0.16 | 0.0004*** | -0.15 ±0.15 | 0.329 | 0.28 ±0.16 | 0.076 |
| Magnetic resonance imaging | |  |  |  |  |  |  |  |  |  |
| Medial Temporal Atrophy | -0.06 ±0.10 | 0.816 | -0.26 ±0.13 | 0.052 | 0.07 ±0.23 | 0.771 | 0.31 ±0.21 | 0.144 | 0.09 ±0.27 | 0.731 |
| Parietal Cortical Atrophy | -0.10 ±0.11 | 0.667 | 0.13 ±0.14 | 0.343 | -0.32 ±0.24 | 0.181 | -0.56 ±0.23 | 0.013* | -0.43 ±0.28 | 0.128 |
| Fazekas score | -0.14 ±0.10 | 0.479 | -0.27 ±0.13 | 0.035* | 0.03 ±0.22 | 0.903 | 0.03 ±0.20 | 0.875 | 0.10 ±0.26 | 0.685 |
| Cerebrospinal fluid |  |  |  |  |  |  |  |  |  |  |
| Aβ42 | -0.09 ±0.10 | 0.667 | -0.04 ±0.13 | 0.760 | 0.21 ±0.20 | 0.297 | -0.64 ±0.21 | 0.003** | -0.18 ±0.25 | 0.475 |
| pTau-181 | 0.03 ±0.10 | 0.847 | 0.00 ±0.13 | 0.994 | 0.60 ±0.20 | 0.003** | -0.50 ±0.21 | 0.019* | -0.04 ±0.25 | 0.877 |
| t-Tau | 0.05 ±0.10 | 0.816 | 0.08 ±0.13 | 0.557 | 0.47 ±0.20 | 0.018* | -0.45 ±0.21 | 0.035* | -0.05 ±0.25 | 0.833 |
| Neurofilament Light | 0.08 ±0.32 | 0.797 | -0.02 ±0.51 | 0.963 | 0.35 ±0.59 | 0.556 | 0.51 ±0.71 | 0.475 | -1.05 ±1.00 | 0.297 |
|  | β ± SE | p-value^✦^ | β ± SE | p-value | β ± SE | p-value | β ± SE | p-value | β ± SE | p-value |
| EEG abnormality^log^ | -0.44 ±0.21 | 0.227 | -0.38 ±0.29 | 0.195 | -0.21 ±0.43 | 0.633 | -0.62 ±0.55 | 0.369 | -0.96 ±0.60 | 0.133 |
| Time diagnosis-death^cox^ | 0.12 ±0.13 | 0.667 | 0.05 ±0.18 | 0.759 | 0.47 ±0.22 | 0.034* | -0.42 ±0.41 | 0.311 | 0.13 ±0.36 | 0.706 |
| Cognitive decline^lmm^ | -0.63 ±0.25 | 0.094^✦^ | -0.54 ±0.31 | 0.085 | -1.40 ±0.61 | 0.021* | -1.75 ±0.74 | 0.018* | 0.58 ±0.62 | 0.350 |

Shown here are *standardized* betas of linear regression models adjusted for age and sex; MMSE at baseline and neuropsychological domains are also adjusted for education level and disease stage. Also shown here are betas of logistic regression models (log), cox proportional hazards models (cox), and linear mixed models (lmm) adjusted for age, sex, education level, and disease stage; cox regression models are also adjusted for MMSE at baseline.

✦ FDR-corrected p<0.10, *p<0.05, **p<0.01.

Other *TREM2* variants include: D87N, G58A, Q33X, C51Y, R47G, R31F, and A105V.

Abbreviations: Aβ42 = Beta-Amyloid 42; CI = confidence interval; EEG = Electroencephalography; EF = Executive Functioning; FDR = False Discovery Rate; MMSE = Mini-Mental State Examination; pTau-181 = Phosphorylated Tau-181; R47H = p.Arg47His; R62H = p.Arg62His; SE = Standard Error; stdβ = standardized beta; T96K = p.Thr96Lys; *TREM2* = Triggering Receptor Expressed On Myeloid Cells 2.

**Supplementary Table 3:** Effect of *TREM2* variants on time between diagnosis and death.

|  | Hazard Ratio [95%-Confidence Interval] | p-value | p-value FDR-corrected |
| --- | --- | --- | --- |
| Model 1 |  |  |  |
| All *TREM2* variants | 1.12 [0.9-1.4] | 0.353 | 0.667 |
| Model 2 |  |  |  |
| R62H | 1.06 [0.8-1.5] | 0.759 | NA |
| R47H | 1.60 [1.0-2.5] | 0.034* | NA |
| T96K | 0.66 [0.3-1.5] | 0.311 | NA |
| Other *TREM2* variants | 1.14 [0.6-2.3] | 0.706 | NA |

Shown here are hazard ratios with 95% confidence interval, adjusted for age at diagnosis, sex, education level, disease stage (MCI/dementia), and MMSE at baseline.

*p<0.05

Other *TREM2* variants include: D87N, G58A, Q33X, C51Y, R47G, R31F, and A105V.

Abbreviations: NA = Not Available; FDR = False Discovery Rate; R47H = p.Arg47His; R62H = p.Arg62His; T96K = p.Thr96Lys; *TREM2* = Triggering Receptor Expressed on Myeloid Cells 2.

**Supplementary Table 4:** Summary statistics of quantitative image analysis of significant *TREM2*-associated cortical thickness regions and subcortical volumes based on Desikan Kiliany atlas.

|  | **Lobe** | **Brain region of interest** | **stdβ ± SE** | **p-value** |
| --- | --- | --- | --- | --- |
| **All *TREM2* variants** | Subcortical | Amygdala | 0.19 ±0.10 | 0.047 |
| **R62H** | Temporal | Temporal pole | 0.29 ±0.14 | 0.037 |
| **R47H** | Occipital | Cuneus | 0.49 ±0.24 | 0.037 |
|  |  | Lingual | 0.47 ±0.24 | 0.049 |
|  | Subcortical | Amygdala | 0.69 ±0.21 | 0.001 |
|  |  | Hippocampus | 0.49 ±0.20 | 0.016 |
| **T96K** | Cingulate | Rostral anterior cingulate | -0.67 ±0.25 | 0.007 |
|  | Frontal | Medial orbitofrontal | -0.50 ±0.25 | 0.047 |
|  | Subcortical | Hippocampus | -0.64 ±0.22 | 0.003 |
|  | NA | Insula | -0.55 ±0.25 | 0.029 |
| **Other *TREM2* variants** | Cingulate | Isthmus cingulate | -0.61 ±0.28 | 0.027 |
|  | Frontal | Pars orbitalis | -0.59 ±0.27 | 0.033 |
|  |  | Frontal pole | -0.56 ±0.28 | 0.042 |

Shown here are standardized betas with standard error, adjusted for age, sex and estimated intracranial volume. The betas represent the effect of *TREM2* variants on cortical thickness (mm) and subcortical volumes (mm^3^) compared to non-carriers with symptomatic AD.

Other *TREM2* variants include: D87N, G58A, Q33X, C51Y, R47G, R31F, and A105V.

Abbreviations: CI = confidence interval; R47H = p.Arg47His; T96K = p.Thr96Lys; SE = Standard Error; stdβ = standardized beta; *TREM2* = Triggering Receptor Expressed on Myeloid Cells 2.

**Supplementary Table 5: Sensitivity analyses of *TREM2* effect on clinical measures in symptomatic Alzheimer’s disease, scaled.**

Shown here are (standardized) betas of the sensitivity analyses. (a) Cohort descriptives per clinical measure. (b) Shown here are standardized betas of linear regression models are adjusted for age, sex, and three principal components. Neuropsychological domains are also adjusted for education level and disease stage (MCI/dementia). Also shown here are betas of logistic regression models (log), cox proportional hazards models (cox), and linear mixed models (lmm) adjusted for age, sex, education level, disease stage and three principal components; cox regression models are also adjusted for MMSE at baseline. (c) Cox regression models are adjusted for age at diagnosis, sex, education level, disease stage, MMSE at baseline, and three principal components. (d) Significant findings of linear regression models in quantitative image analysis adjusted for age, sex, estimated intracranial volume, and three principal components (p<0.05).

✦ FDR-corrected p<0.10, *p<0.05, **p<0.01, ***p<0.001.

Other *TREM2* variants include: D87N, G58A, Q33X, C51Y, R31F.

Abbreviations: Aβ42 = Beta-Amyloid 42; AD = Alzheimer’s Disease; ApoE = Apolipoprotein E; CI = confidence interval; CSF = cerebrospinal fluid; EF = Executive Functioning; IQR = Interquartile Range; MMSE = Mini-Mental State Examination; MRI = Magnetic Resonance Imaging; MTA = Medial Temporal Lobe; *n* = number; NA = Not Applicable; NfL = Neurofilament Light; Neuropsych. = Neuropsychological; PCA = Posterior Cortical Atrophy; pTau-181 = Phosphorylated Tau-181; R47H = p.Arg47His; R62H = p.Arg62His; SD = Standard Deviation; SE = Standard Error; stdβ = standardized beta; T96K = p.Thr96Lys; *TREM2* = Triggering Receptor Expressed on Myeloid Cells 2.

a)

|  | **Main analysis** | |  |  |  |  |  |  | **Exploratory analysis** | |
| --- | --- | --- | --- | --- | --- | --- | --- | --- | --- | --- |
|  | Total | Neuropsych.  Domains | MRI clinical  ratings | CSF | CSF-NfL | EEG | Time Diagnosis  to Death | MMSE  over Time | MRI structural | Tau-PET |
| Total | 1444 (100) | 1391 (96) | 1102 (76) | 1390 (96) | 150 (10) | 1189 (82) | 1444 (100) | 1431 (99) | 968 (67) | 63 (4) |
| Female | 747 (52) | 717 (52) | 572 (52) | 719 (52) | 85 (57) | 616 (52) | 747 (52) | 739 (52) | 496 (51) | 31 (49) |
| *TREM2* mutation carriers | 103 (7) | 101 (7) | 84 (8) | 97 (7) | 8 (5) | 89 (7) | 103 (7) | 101 (7) | 83 (9) | 4 (6) |
| R62H | 60 (58) | 59 (58) | 51 (61) | 55 (57) | 4 (50) | 52 (58) | 60 (58) | 60 (59) | 50 (60) | 1 (25) |
| R47H | 26 (25) | 25 (25) | 18 (21) | 26 (27) | 3 (38) | 23 (26) | 26 (25) | 24 (24) | 18 (22) | 1 (25) |
| T96K | 3 (3) | 3 (3) | 3 (4) | 3 (3) | 0 (0) | 2 (2) | 3 (3) | 3 (3) | 2 (2) | 0 (0) |
| Other variants* | 16 (16) | 16 (16) | 13 (15) | 15 (15) | 1 (12) | 14 (16) | 16 (16) | 16 (16) | 14 (17) | 2 (50) |

Table shows *n* (%).

b)

|  | **Main analysis** | | **Exploratory analysis** | |  |  |  |  |  | |
| --- | --- | --- | --- | --- | --- | --- | --- | --- | --- | --- |
|  | All *TREM2* variants combined | | R62H |  | R47H |  | T96K |  | Other *TREM2* variants | |
|  | stdβ ± SE | p-value^✦^ | stdβ ± SE | p-value | stdβ ± SE | p-value | stdβ ± SE | p-value | stdβ ± SE | p-value |
| MMSE at baseline | 0.09 ±0.10 | 0.555 | 0.08 ±0.13 | 0.506 | -0.14 ±0.20 | 0.497 | 0.45 ±0.56 | 0.419 | 0.31 ±0.24 | 0.196 |
| Neuropsychological domains | |  |  |  |  |  |  |  |  |  |
| Memory | -0.08 ±0.08 | 0.555 | -0.08 ±0.10 | 0.443 | -0.29 ±0.17 | 0.079 | -0.43 ±0.45 | 0.340 | 0.22 ±0.20 | 0.270 |
| Executive functioning | 0.16 ±0.17 | 0.555 | 0.26 ±0.22 | 0.233 | -0.69 ±0.35 | 0.052 | -0.46 ±0.91 | 0.612 | 0.81 ±0.40 | 0.046* |
| Attention and speed | 0.08 ±0.11 | 0.700 | 0.25 ±0.14 | 0.082 | -0.40 ±0.22 | 0.072 | 0.58 ±0.60 | 0.337 | 0.06 ±0.26 | 0.832 |
| Language | -0.01 ±0.08 | 0.943 | 0.15 ±0.10 | 0.153 | -0.39 ±0.17 | 0.022* | -0.36 ±0.45 | 0.430 | 0.02 ±0.20 | 0.904 |
| Visuospatial functioning | 0.22 ±0.08 | 0.136 | 0.19 ±0.11 | 0.072 | 0.23 ±0.17 | 0.188 | 0.20 ±0.41 | 0.629 | 0.30 ±0.19 | 0.122 |
| Global cognition | -0.03 ±0.07 | 0.872 | 0.03 ±0.09 | 0.765 | -0.56 ±0.16 | 0.0004*** | -0.18 ±0.34 | 0.589 | 0.27 ±0.16 | 0.082 |
| Magnetic resonance imaging | |  |  |  |  |  |  |  |  |  |
| Medial Temporal Atrophy | -0.12 ±0.11 | 0.555 | -0.23 ±0.14 | 0.099 | 0.06 ±0.23 | 0.783 | 0.00 ±0.56 | 0.998 | 0.08 ±0.27 | 0.770 |
| Parietal Cortical Atrophy | -0.02 ±0.11 | 0.943 | 0.19 ±0.14 | 0.189 | -0.34 ±0.24 | 0.150 | -0.45 ±0.71 | 0.525 | -0.44 ±0.28 | 0.116 |
| Fazekas score | -0.18 ±0.11 | 0.353 | -0.30 ±0.13 | 0.025* | 0.02 ±0.22 | 0.914 | -0.58 ±0.54 | 0.281 | 0.08 ±0.26 | 0.755 |
| Cerebrospinal fluid |  |  |  |  |  |  |  |  |  |  |
| Aβ42 | 0.02 ±0.10 | 0.943 | -0.00 ±0.14 | 0.994 | 0.22 ±0.20 | 0.273 | -0.52 ±0.57 | 0.365 | -0.17 ±0.26 | 0.498 |
| pTau-181 | 0.11 ±0.10 | 0.555 | -0.00 ±0.14 | 0.989 | 0.60 ±0.20 | 0.003** | -0.63 ±0.58 | 0.276 | -0.05 ±0.26 | 0.839 |
| t-Tau | 0.11 ±0.10 | 0.555 | 0.08 ±0.14 | 0.548 | 0.46 ±0.20 | 0.020* | -0.74 ±0.58 | 0.199 | -0.09 ±0.26 | 0.735 |
| Neurofilament Light | -0.06 ±0.37 | 0.865 | -0.04 ±0.52 | 0.931 | 0.28 ±0.60 | 0.638 | NA | NA | NA | NA |
|  | β ± SE | p-value^✦^ | β ± SE | p-value | β ± SE | p-value | β ± SE | p-value | β ± SE | p-value |
| EEG abnormality^log^ | -0.49 ±0.23 | 0.249 | -0.37 ±0.30 | 0.205 | -0.21 ±0.43 | 0.631 | -12.9 ±NA | 0.969 | -1.2 ±0.67 | 0.065 |
| Time diagnosis-death^cox^ | 0.14 ±0.13 | 0.555 | -0.01 ±0.18 | 0.951 | 0.46 ±0.22 | 0.038* | 0.08 ±1.0 | 0.934 | 0.08 ±0.38 | 0.844 |
| Cognitive decline^lmm^ | -0.52 ±0.26 | 0.249 | -0.61 ±0.32 | 0.054 | -1.38 ±0.61 | 0.022* | 0.26 ±1.2 | 0.833 | 0.61 ±0.63 | 0.332 |

c)

|  | Hazard Ratio [95% Confidence Interval] | p-value | p-value FDR-corrected |
| --- | --- | --- | --- |
| All *TREM2* variants | 1.15 [0.9-1.5] | 0.289 | 0.555 |
| R62H | 0.99 [0.7-1.4] | 0.951 | NA |
| R47H | 1.59 [1.0-2.5] | 0.038* | NA |
| T96K | 1.09 [0.1-7.8] | 0.934 | NA |
| Other *TREM2* variants | 1.08 [0.5-2.3] | 0.844 | NA |

d)

|  | Lobe | Brain region of interest | stdβ ± SE | p-value |
| --- | --- | --- | --- | --- |
| All *TREM2* variants | Subcortical | Amygdala | 0.21 ±0.10 | 0.039 |
| R47H | Occipital | Cuneus | 0.48 ±0.24 | 0.041 |
|  | Subcortical | Amygdala | 0.70 ±0.21 | 0.001 |
|  |  | Hippocampus | 0.47 ±0.20 | 0.023 |
| T96K | Temporal | Parahippocampal | 1.55 ±0.69 | 0.025 |
| Other *TREM2* variants | Cingulate | Isthmus cingulate | -0.61 ±0.28 | 0.028 |
|  | Frontal | Pars orbitalis | -0.55 ±0.28 | 0.046 |
